# Suicide Risk of Third-Generation Antipsychotics in Persons with Schizophrenia and Schizoaffective Disorders: A Systematic Review and Meta-Analysis

**DOI:** 10.64898/2026.02.10.26345876

**Authors:** Jeff W. Jin, Charlotte J. Winkler, Heather B. Blunt, Natalie B. Riblet

## Abstract

**Background and Hypothesis:** Clozapine is the only antipsychotic with protective effects against suicide in schizophrenia (SCZ). Newer third-generation antipsychotics (TGA) have better tolerability and modulate serotonin, dopamine, and N-methyl-d-aspartate neurotransmission pathways implicated in suicide. We aimed to investigate the effects of TGAs on suicide in SCZ.

**Methods:** We searched seven databases up to December 2023 for SCZ studies that reported suicide data. The primary outcome was suicide deaths and attempts; suicidal ideation was added as a secondary outcome. Random effects meta-analyses quantified suicide risk in randomized controlled trials (RCT) while single proportion meta-analyses assessed longitudinal suicide risk in open label extension trials (OLE). For RCTs, sensitivity analyses were conducted and subgroup analyses explored the impact of dose, drug type, and comparator arm.

**Study Results:** Twenty articles were included; thirteen excluded higher suicide risk participants. Compared to placebo control, TGAs did not significantly change the risk of primary [RR = 0.65, p = 0.38] or secondary [RR = 0.63, p = 0.15] suicide outcomes. Subgroup and sensitivity analyses were not statistically significant. For OLEs, there was a significant increase in the incidence of primary [Ip = 0.004, p = 0.048] and secondary [Ip = 0.024, p = 0.0013] suicide outcomes, but there was marked study heterogeneity.

**Conclusion:** There is no current trial evidence to show that TGAs significantly impact suicide outcomes in SCZ. The signal from OLEs should be interpreted cautiously due to heterogeneity and requires replication. An effective clozapine alternative is needed for suicide prevention in SCZ.

## 1. Introduction

Schizophrenia spectrum disorders (SCZ) primarily include schizophreniform, schizophrenia, and schizoaffective disorders and have an estimated lifetime prevalence of 1%.^1, 2^ SCZ is a significant contributor to the global burden of disease and has increased in burden contribution over time.^3, 4^ In the United States in 2019, the estimated cost burden of SCZ was $343.2 billion with substantial indirect costs from premature mortality.^5^ There is a substantial mortality gap between persons with schizophrenia and the general population.^6^ Reductions in life expectancy in SCZ are estimated between 15–20 years with high contributions from cardiometabolic disease and suicide.^6, 7^

Suicide is a leading cause of premature death in SCZ.^8^ The lifetime world prevalence of suicide attempts in SCZ is 26.8% with a significant increased prevalence in North America (35.9%) and Europe and Central Asia (32.2%).^9^ Around 4–10% of persons with SCZ will die by suicide during their lifetimes, with male sex, younger age, and higher intelligence as significant risk factors.^10-12^ More broadly, suicidal behavior has been linked to serotonergic dysfunction through alterations in cerebrospinal fluid serotonin metabolites and post-mortem serotonin neurotransmission.^13, 14^ Additionally, studies suggest a relationship between suicidal behaviors and dopaminergic dysfunction, a hypothesized etiology of SCZ, due to dysregulation in striatal dopamine receptor binding and predictability of suicidal ideation from reduced dorsal striatal gray matter volume.^15-17^ Furthermore, *N*-methyl-d-aspartate (NMDA) receptor alterations observed in the frontal cortex and hippocampus in suicide victims suggest implications of NMDA dysfunction in suicide.^18, 19^

Despite high occurrence of suicide in SCZ, pharmacologic interventions specifically targeting suicide remain very limited. Clozapine is the only antipsychotic with protective effects against suicide in SCZ.^20, 21^ Additionally, clozapine demonstrates superior efficacy in the reduction of positive and negative symptoms, overall psychopathology, and relapse rates in SCZ compared to first and second-generation antipsychotics.^22^ However, the efficacy of clozapine is countered by shifting monitoring protocols and significant side effects including neutropenia, constipation, hypotension, sedation, and weight gain that pose additional barriers to existing adherence difficulties for persons with SCZ.^23-25^

Third-generation antipsychotics (TGA) primarily include aripiprazole, oxaripiprazole, brexpiprazole, cariprazine, and lumateperone and are a newer medication class characterized by their unique modulation of dopamine through partial agonism of D2 and D3 receptors.^26, 27^ Compared to other antipsychotics, TGAs have increased tolerability due to lower risks of weight gain, hyperprolactinemia, sedation, and anticholinergic side effects which are associated with low discontinuation rates.^27^ Contrary to clozapine, TGA-associated lab monitoring is much less rigorous and follows standard antipsychotic monitoring guidelines. Of particular relevance to suicide prevention, several TGAs also modulate serotonin activity through potent serotonin transporter (SERT) inhibition, 5-hydroxytryptamine (5-HT) 1A receptor partial agonism, and 5-HT2A and 2B receptor antagonism.^28-30^ Additionally, lumateperone enhances NMDA receptor function and indirectly modulates glutamine neurotransmission.^31^

Suicide in SCZ has considerable negative burdens on quality of life, financial costs, and premature mortality. Pharmacologic interventions for suicide in SCZ are limited to clozapine, an effective antipsychotic with notable burdens of lab monitoring and side effects. There is a clear need for a more benign antipsychotic alternative with protective effects against suicide. TGAs may fill this gap because of pharmacologic activity in neurotransmitter pathways implicated in suicidal behavior. Given this, we conducted a systematic review and meta-analysis to determine the effects of TGAs on suicide risk in persons with SCZ.

## 2. Methods

For the current study, we registered the protocol (CRD42023449336) with the International Prospective Register of Systematic Reviews (PROSPERO) prior to conducting the final data analysis and followed the Preferred Reporting Items for Systematic Reviews and Meta-Analyses (PRISMA) guidelines.^32, 33^

### 2.1 Search Strategy

We conducted a search of PubMed, MEDLINE, PsycINFO, Scopus, Cochrane Central Register of Controlled Trials, Europe PMC, and ClinicalTrials.gov databases from inception up to December 2023. The search strategy focused on medical subject headings (MeSH) and variations of keywords related to: (“Schizophrenia”) AND (“Suicide”) AND (“Antipsychotic”) AND (“Clinical Trial”). The search strategy was designed and peer-reviewed by two independent librarians and search terms were adapted to respective databases. Please refer to the supplementary materials for further detailed information on search strategy.

### 2.2 Inclusion and Exclusion Criteria

The major inclusion criteria were: [1] publication in English language, [2] studies of human subjects, [3] randomized controlled trials (RCT) or open-label extension studies (OLE) of completed randomized controlled trials, [4] inclusion of individuals with Diagnostic and Statistical Manual of Mental Disorders, Fourth Edition (DSM-IV), or revised diagnosis of SCZ (schizophrenia, schizoaffective disorder, schizophreniform disorder), [5] intervention of third-generation antipsychotic agents including brexpiprazole, cariprazine, lumateperone, and oxaripiprazole, and [6] reporting of outcomes or suicide deaths, attempts, or ideation.

The notable exclusion criteria were: [1] non-SCZ conditions or diagnoses including subclinical or prodromal states (early phase psychosis, clinical high-risk for psychosis), substance-induced psychosis, delusional disorder, or schizotypal personality disorder, [2] non-TGA interventions and aripiprazole, [3] lack of reported suicide outcomes (deaths, attempts, or ideation) or studies where data on suicide outcomes were not able to be obtained, and [4] open-label trials that were not extensions of or did not include participants from prior RCTs. Studies on aripiprazole were excluded to focus on newer third-generation antipsychotic agents. Articles without specific report of suicide outcomes were excluded if data on suicide outcomes were not obtainable from supplementary materials, results from clinical trials registries, or contact with the corresponding author or listed trial contact or sponsor.

### 2.3 Screening and Data Extraction

Two reviewers (JJ, NR) independently screened the search results by title, abstract, full manuscript (where indicated), and eligibility using Rayyan QCRI software.^34^ Disagreements were resolved through discussion and consensus agreement. Two independent reviewers (JJ, CW) extracted data from eligible studies for formal analysis using a standardized data collection form.

### 2.4 Quality Assessment

Two reviewers (JJ, NR) independently assessed study quality and bias risk assessment with the revised Cochrane Risk of Bias Tool (RoB 2) across five domains: randomization, intervention assignment and adherence, missing outcome data, outcome measurement, and selection or reported results.^35^ The overall bias risk for all studies was determined to be low risk, some bias concerns, or high risk. Disagreements were resolved through discussion and consensus agreement.

### 2.5 Outcomes

Primary outcomes included suicide deaths and attempts. We chose these measures since they are the most robust measures of suicide risk. Secondary outcomes included suicide deaths, attempts, and ideation. Suicidal ideation was included as a secondary outcome as suicide deaths and attempts are a rare occurrence that may be further limited by the short-term duration of randomized controlled trials and exclusion of participants with higher suicide risk.

### 2.6 Data Synthesis and Analysis

We conducted random effects meta-analyses for RCTs and single proportion meta-analyses for single-arm OLEs of suicide outcomes using Stata SE18 software. Statistical significance was measured at *p* < 0.05. Risk ratios (RR) and 95% confidence intervals (95% CI) were calculated to estimate intervention effect size. For study arms with zero events, a continuity correction of 0.5 was added to prevent mathematical impossibility.^36, 37^ The Peto Odds Ratio was considered due to the presence of arms with zero outcome events, but was ultimately not used because of the risks of effect overestimation due to unbalanced group sizes.^38^ Heterogeneity was measured using *I*^2^, *H*^2^, and Cochran Q test statistics. Heterogeneity was defined as an *I*^2^ > 50% and a *p* < 0.1. Meta-analysis results were presented in forest plots. Publications bias was assessed with funnel plots and Egger’s tests. For RCTs, sensitivity analyses were conducted to exclude studies classified as having “some concerns” or “high risk” of bias.

Additionally, subgroups analyses were conducted to explore the impact of dose, drug type, and choice of comparator arm.

## 3. Results

### 3.1 Search Results and Quality Assessment

The initial search resulted in 4418 articles and 3063 articles remained after removal of 1355 duplicates. After the exclusion of 2728 articles from abstract screening, 335 articles were retrieved for full review. After article review, 20 articles met inclusion criteria (Supplementary Figure S1) including 15 RCTs (*N* = 5700) – five brexpiprazole (*N* = 2260), seven cariprazine (*N* = 2703), two lumateperone (*N* = 701), and one oxaripiprazole (*N* = 36). Five OLEs (*N* = 2200) also met inclusion criteria – three brexpiprazole (*N* = 1521) and two cariprazine (*N* = 679).

Detailed characteristics of the included studies are summarized in Table 1. Study samples ranged from 32 to 729 participants for RCTs and 93 to 1031 participants for OLEs. Brexpiprazole and cariprazine were the most studied TGAs. 11 RCTs had a study duration of 6 weeks or less while all OLEs have trial periods of around 1 year. The Columbia Suicide Severity Rating Scale (C-SSRS) was the most common method of suicide assessment; 13 studies excluded participants with increased risk of suicide. For comparator arms in RCTs, 14 RCTs used a placebo control condition while two of these RCTs had additional aripiprazole and quetiapine arms. One RCT used risperidone without a control condition. Quality assessment showed 13 RCTs with low bias risk and two RCTs with some concerns of bias. All five OLEs had some concerns of bias. Risk of bias assessments are summarized in supplementary materials (Figures S2, S3; Tables S1, S2).

**Table 1.**
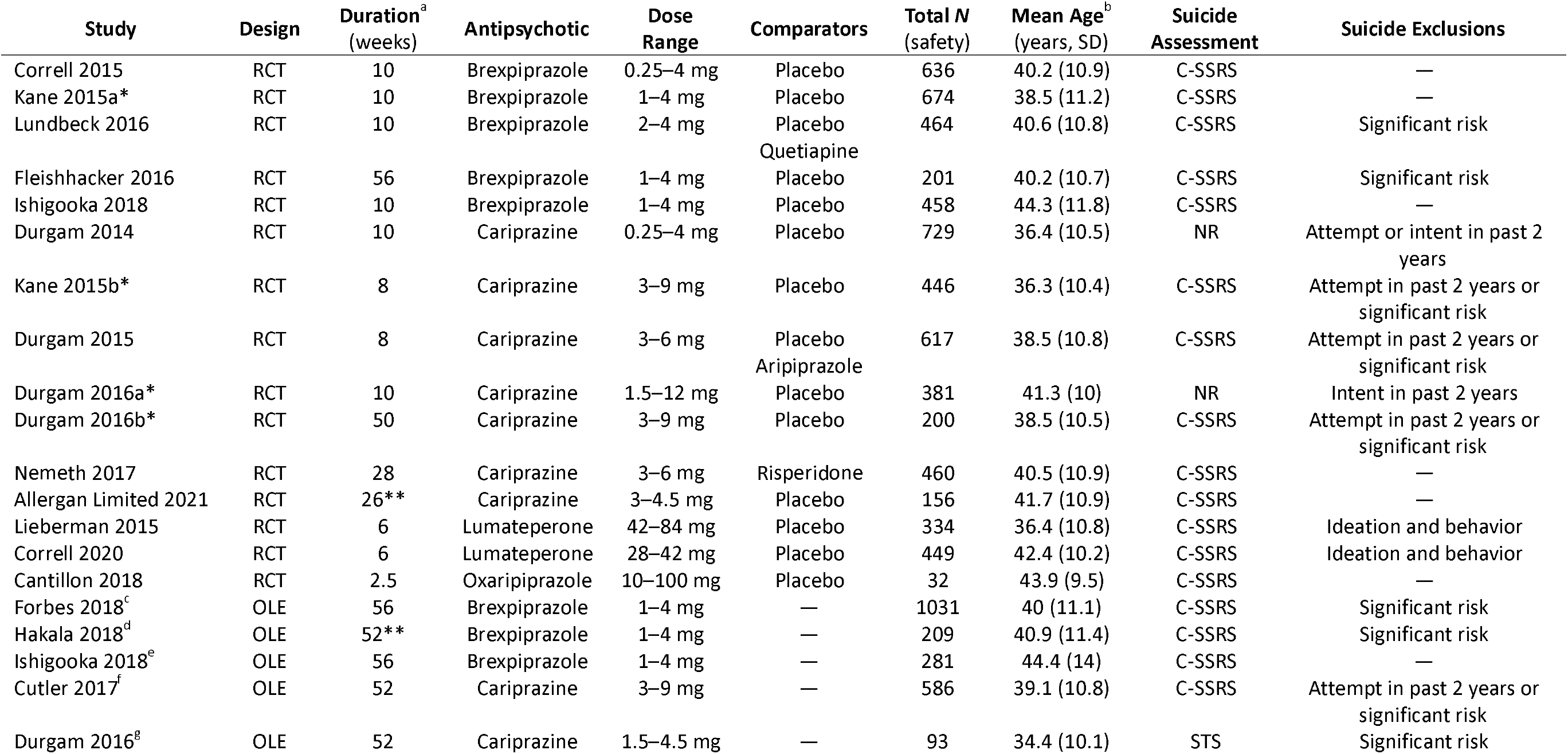
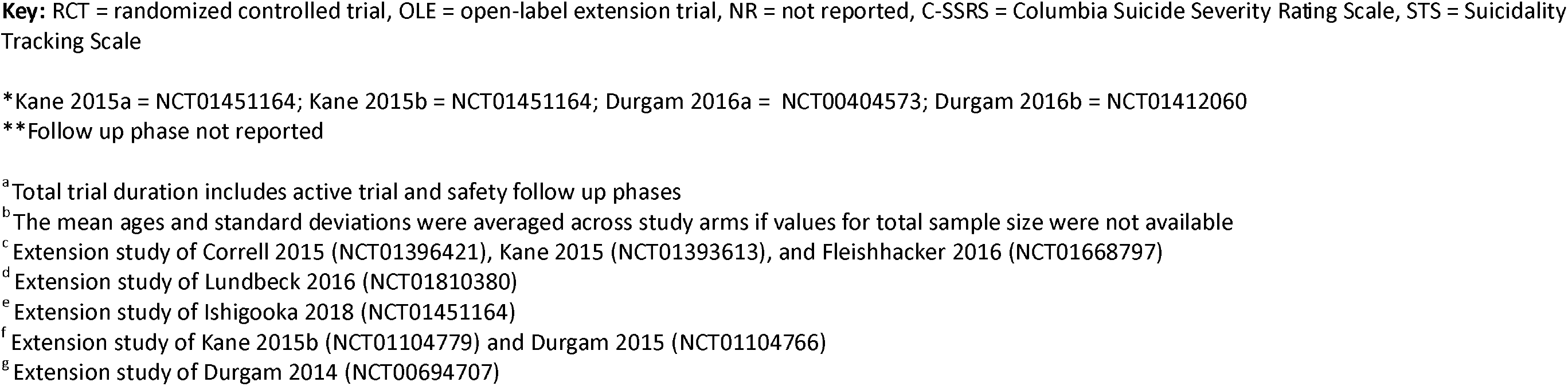
Characteristics of Included Randomized Controlled Trials and Open-label Extension Trials.

| Study | Design | Duration<sup>a</sup><br>(weeks) | Antipsychotic | Dose<br>Range | Comparators | Total N<br>(safety) | Mean Age<sup>b</sup><br>(years, SD) | Suicide<br>Assessment | Suicide Exclusions |
| --- | --- | --- | --- | --- | --- | --- | --- | --- | --- |
| Correll 2015 | RCT | 10 | Brexpiprazole | 0.25–4 mg | Placebo | 636 | 40.2 (10.9) | C-SSRS | — |
| Kane 2015a* | RCT | 10 | Brexpiprazole | 1–4 mg | Placebo | 674 | 38.5 (11.2) | C-SSRS | — |
| Lundbeck 2016 | RCT | 10 | Brexpiprazole | 2–4 mg | Placebo | 464 | 40.6 (10.8) | C-SSRS | Significant risk |
|  |  |  |  |  | Quetiapine |  |  |  |  |
| Fleishhacker 2016 | RCT | 56 | Brexpiprazole | 1–4 mg | Placebo | 201 | 40.2 (10.7) | C-SSRS | Significant risk |
| Ishigooka 2018 | RCT | 10 | Brexpiprazole | 1–4 mg | Placebo | 458 | 44.3 (11.8) | C-SSRS | — |
| Durgam 2014 | RCT | 10 | Cariprazine | 0.25–4 mg | Placebo | 729 | 36.4 (10.5) | NR | Attempt or intent in past 2 years |
| Kane 2015b* | RCT | 8 | Cariprazine | 3–9 mg | Placebo | 446 | 36.3 (10.4) | C-SSRS | Attempt in past 2 years or significant risk |
| Durgam 2015 | RCT | 8 | Cariprazine | 3–6 mg | Placebo | 617 | 38.5 (10.8) | C-SSRS | Attempt in past 2 years or significant risk |
|  |  |  |  |  | Aripiprazole |  |  |  |  |
| Durgam 2016a* | RCT | 10 | Cariprazine | 1.5–12 mg | Placebo | 381 | 41.3 (10) | NR | Intent in past 2 years |
| Durgam 2016b* | RCT | 50 | Cariprazine | 3–9 mg | Placebo | 200 | 38.5 (10.5) | C-SSRS | Attempt in past 2 years or significant risk |
| Nemeth 2017 | RCT | 28 | Cariprazine | 3–6 mg | Risperidone | 460 | 40.5 (10.9) | C-SSRS | — |
| Allergan Limited 2021 | RCT | 26** | Cariprazine | 3–4.5 mg | Placebo | 156 | 41.7 (10.9) | C-SSRS | — |
| Lieberman 2015 | RCT | 6 | Lumateperone | 42–84 mg | Placebo | 334 | 36.4 (10.8) | C-SSRS | Ideation and behavior |
| Correll 2020 | RCT | 6 | Lumateperone | 28–42 mg | Placebo | 449 | 42.4 (10.2) | C-SSRS | Ideation and behavior |
| Cantillon 2018 | RCT | 2.5 | Oxaripiprazole | 10–100 mg | Placebo | 32 | 43.9 (9.5) | C-SSRS | — |
| Forbes 2018<sup>c</sup> | OLE | 56 | Brexpiprazole | 1–4 mg | — | 1031 | 40 (11.1) | C-SSRS | Significant risk |
| Hakala 2018<sup>d</sup> | OLE | 52** | Brexpiprazole | 1–4 mg | — | 209 | 40.9 (11.4) | C-SSRS | Significant risk |
| Ishigooka 2018<sup>e</sup> | OLE | 56 | Brexpiprazole | 1–4 mg | — | 281 | 44.4 (14) | C-SSRS | — |
| Cutler 2017<sup>f</sup> | OLE | 52 | Cariprazine | 3–9 mg | — | 586 | 39.1 (10.8) | C-SSRS | Attempt in past 2 years or significant risk |
| Durgam 2016<sup>g</sup> | OLE | 52 | Cariprazine | 1.5–4.5 mg | — | 93 | 34.4 (10.1) | STS | Significant risk |

**Key:** RCT = randomized controlled trial, OLE = open-label extension trial, NR = not reported, C-SSRS = Columbia Suicide Severity Rating Scale, STS = Suicidality Tracking Scale
\*Kane 2015a = NCT01451164; Kane 2015b = NCT01451164; Durgam 2016a = NCT00404573; Durgam 2016b = NCT01412060
\*\*Follow up phase not reported
<sup>a</sup> Total trial duration includes active trial and safety follow up phases
<sup>b</sup> The mean ages and standard deviations were averaged across study arms if values for total sample size were not available
<sup>c</sup> Extension study of Correll 2015 (NCT01396421), Kane 2015 (NCT01393613), and Fleishhacker 2016 (NCT01668797)
<sup>d</sup> Extension study of Lundbeck 2016 (NCT01810380)
<sup>e</sup> Extension study of Ishigooka 2018 (NCT01451164)
<sup>f</sup> Extension study of Kane 2015b (NCT01104779) and Durgam 2015 (NCT01104766)

### 3.2 Meta-analysis of Randomized Controlled Trials

Of the 14 RCTs comparing TGAs to placebo control, there was no significant change in the risk of suicide deaths or attempts [RR = 0.65, 95% CI = 0.24–1.72, *p* = 0.38] and low concern for heterogeneity [*I*^2^ = 0%, *H*^2^ = 1; Q(13) = 1.88, *p* = 1] (Figure 1). There was also no significant change in the risk of suicide deaths, attempts, or ideation [RR = 0.63, 95% CI = 0.34–1.19, *p* = 0.15] and low concern for heterogeneity [*I*^2^ = 0%, *H*^2^ = 1; Q(13) = 2.92, *p* = 1] (Figure 2). Sensitivity analyses of both primary and secondary suicide outcomes did not yield statistically different results (Supplementary Figures S4, S5).

**Figure 1.**
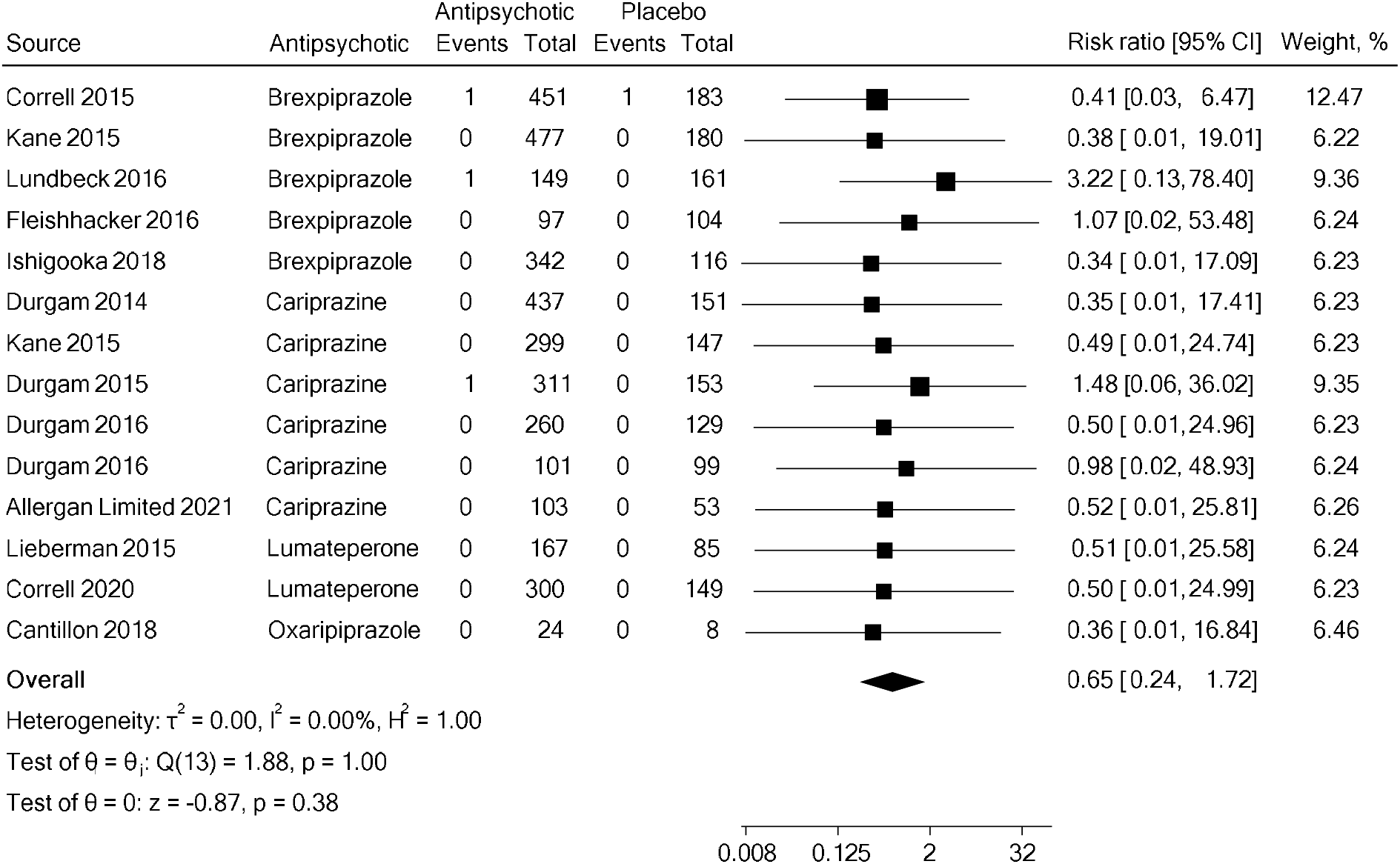
Meta-analysis of Suicide Deaths and Attempts in Randomized Controlled trials

**Figure 2.**
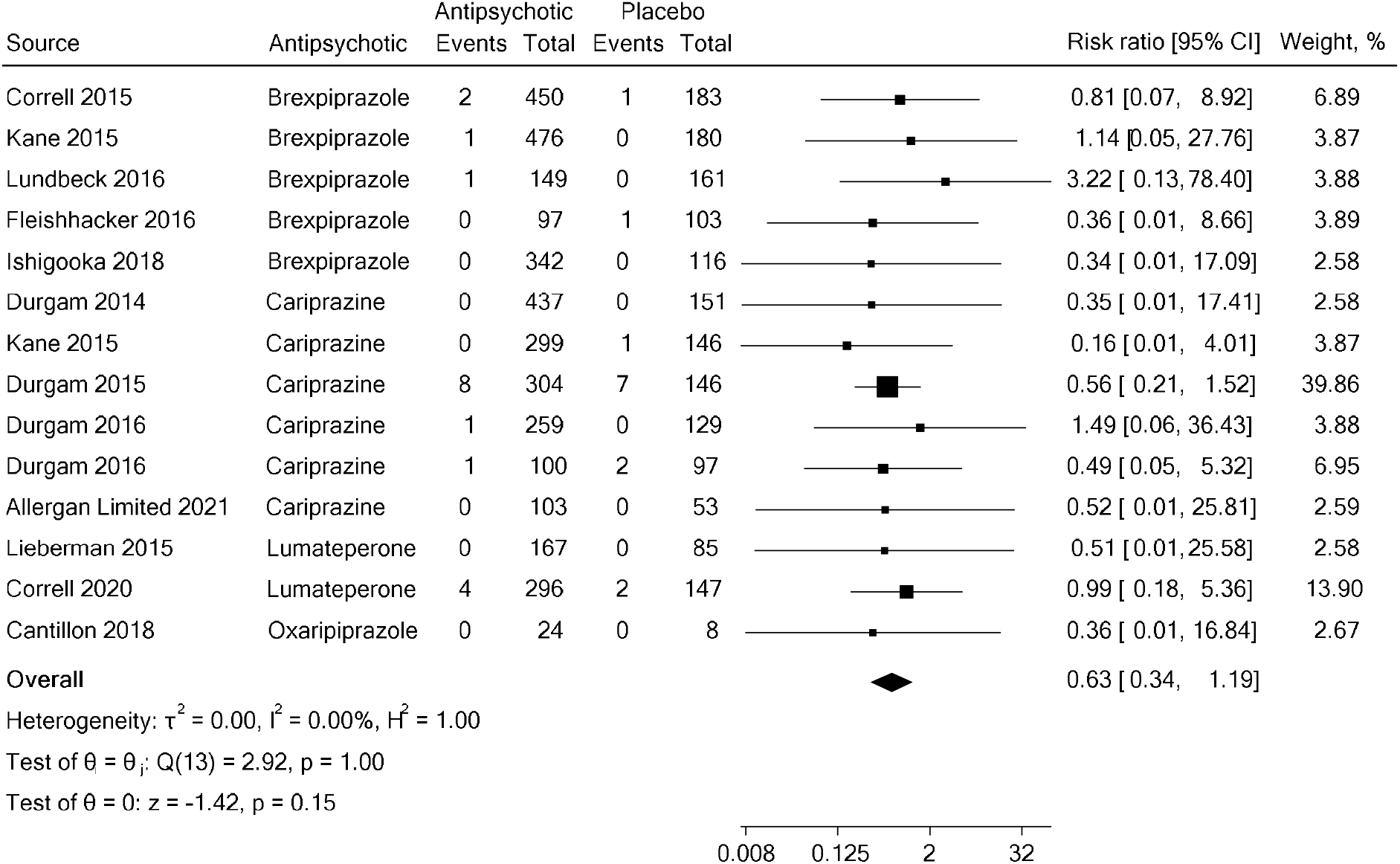
Meta-analysis of Suicide Deaths, Attempts, and Ideation in Randomized Controlled trials

Subgroups analyses by antipsychotic agent (brexpiprazole, cariprazine), antipsychotic dose (brexpiprazole < 2mg, brexpiprazole ≥ 2 mg, cariprazine < 6 mg, cariprazine ≥ 6 mg), and choice of comparator arm (alternative antipsychotics of aripiprazole, quetiapine, and risperidone) did not reveal significant changes in primary or secondary suicide outcomes (Supplementary Figures S6, S8, S10, S12, S14, S16). There were low concerns for heterogeneity across all subgroup analyses. Sensitivity analyses showed no additional changes in significance (Supplementary Figures S7, S9, S11, S13, S15, S17). Further details on subgroup analyses are provided in the supplementary materials.

### 3.3 Meta-analysis of Open-Label Extension Trials

Single proportion meta-analysis of OLEs that followed patients between 48–52 weeks showed a small but significant increase in the incidence of suicide attempts and deaths [pooled incidence = 0.004, 95% CI = 0.000–0.013, *p* = 0.048] (Figure 3) and suicide deaths, attempts, and ideation [pooled incidence = 0.024, 95% CI = 0.005–0.054, *p* + 0.0013] (Figure 4); however, there was marked heterogeneity across primary [*I*^2^ = 71.68%, *H*^2^ = 3.53; Q(4) = 12.75, *p* = 0.013] and secondary [*I*^2^ = 91.32%, *H*^2^ = 11.52; Q(4) = 55.50, *p* < 0.0001] suicide outcomes.

**Figure 3.**
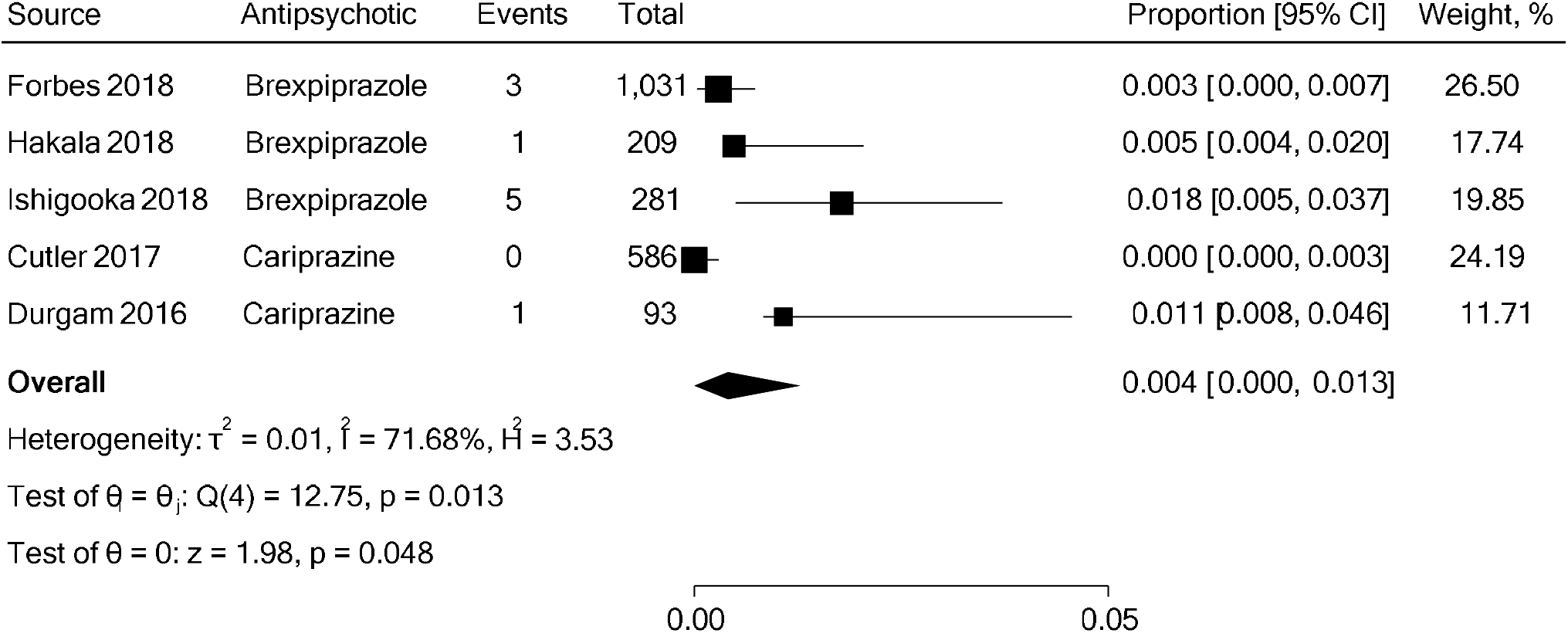
Single Proportion Meta-analysis of Suicide Deaths and Attempts in Open-label Extension trials

**Figure 4.**
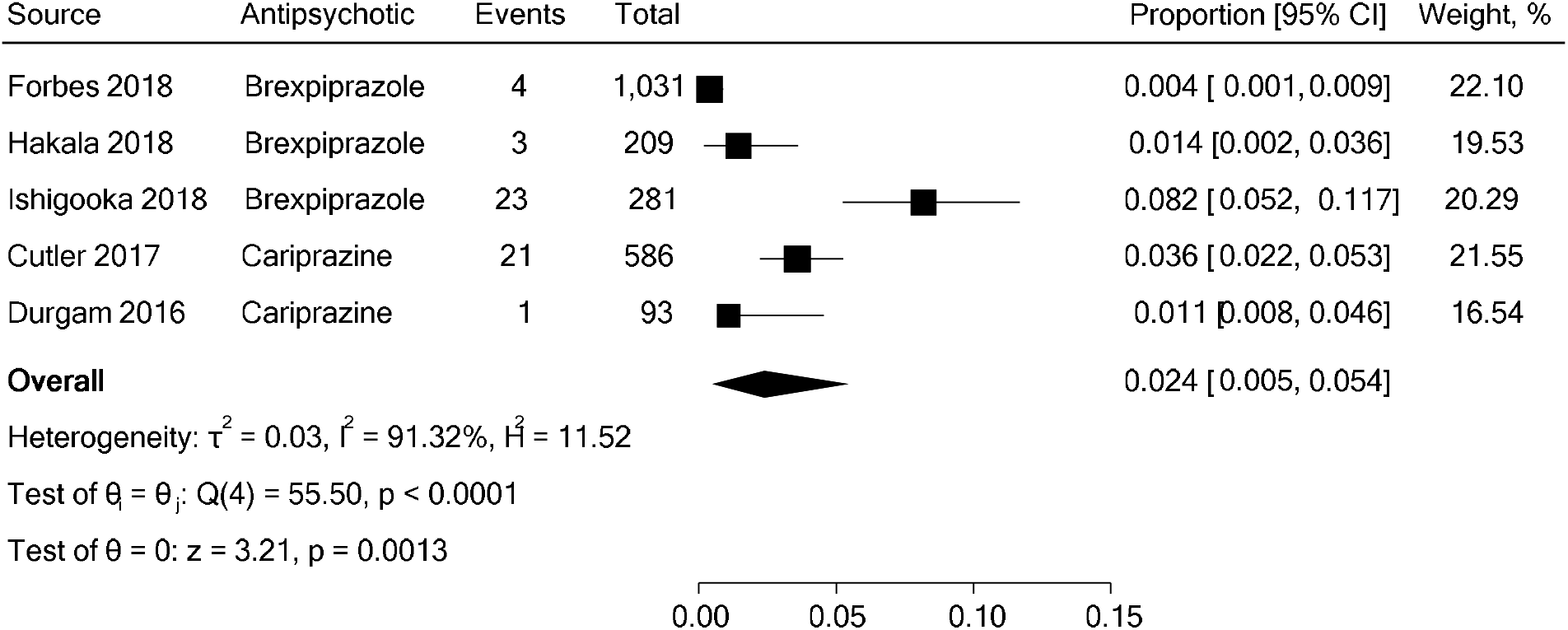
Single Proportion Meta-analysis of Suicide Deaths, Attempts, and Ideation in Open-label Extension trials

### 3.4 Publication Bias

The constructed funnel plots demonstrated no qualitative evidence of asymmetry and was confirmed with Egger’s regression test for small-study effects for RCTs [*t* = -1.20, *p* = 0.25] and OLEs [*t* = 1.35, *p* = 0.27], suggesting no substantial publication bias across included studies (Supplementary Figures S18, S19).

## 4. Discussion

Our systematic review and meta-analysis of 15 RCTs with 5700 total participants with SCZ showed no significant effect of TGAs on primary and secondary suicide outcomes compared to placebo control. Additionally, subgroup analyses by antipsychotic agent, antipsychotic dose, and choice of comparator arm showed no significant effect on suicide outcomes. Single proportion meta-analysis of OLEs of brexpiprazole and cariprazine with 2200 total participants with SCZ showed a statistically significant increase of primary and secondary suicide outcomes, however, there was high heterogeneity between studies. Sensitivity analyses for RCTs did not yield statistical differences across all study results.

Among the included RCTs, there was no significant effect of TGAs on suicide risk. The rarity of suicide, short-term trial durations, and intentional exclusion of patients with higher suicide risk may have limited the occurrence and capture of suicide events. Patients at high risk of suicide were also typically excluded from trials. No increased suicide risk was observed in subgroup analysis of brexpiprazole and cariprazine, agents with higher akathisia risk, by agent and dose.^27^ However, in OLEs investigating moderate doses of brexpiprazole and cariprazine, there was a significantly increased pooled incidence of suicide events, but high heterogeneity across studies limits the conclusiveness of these findings. Our findings are consistent with prior research and demonstrate no current evidence to support a linkage between akathisia and suicidality, but a possible increased signal influenced by high heterogeneity that needs replication.^39-43^ One non-extension open-label study of brexpiprazole and another open-label study phase of cariprazine also showed a high amount of suicide deaths, attempts, and ideation, consistent with our findings.^44, 45^ This suggests that longer trial periods may be necessary to adequately observe and evaluate suicide risk.

Only two RCTs for lumateperone were included and both trials had no suicide attempts or deaths in either study arm.^46, 47^ There were an inadequate number of studies to conduct a subgroup analysis of the effects of lumateperone on suicide outcomes. Additionally, there were no eligible OLEs for lumateperone to describe longitudinal effects on suicide outcomes. The low number of studies and suicide events limits our ability to comment on the impact of lumateperone on suicide risk in SCZ. However, the potential for lumateperone to impact suicide remains interesting due to its enhancement of NMDA receptors that are implicated in suicide.^18, 19^

Our findings further support the unique protective effect of clozapine against suicide in SCZ. This is consistent with the International Suicide Prevention Trial (InterSePT) where clozapine was superior to olanzapine in prevention of suicide attempts.^20^ A prior meta-analysis including additional registry studies also observed an association between clozapine and reduction of suicidal behavior;^21^ however, there are notable concerns from the inclusion of non-randomized studies with varying methodologies that may compound effects from flaws in study design and confounding variables.^48^ Additional research also shows a large and consistent effect of clozapine on suicidal risk and lack thereof from alternative antipsychotic agents.^49^

Most of the eligible studies excluded participants that were at a higher risk for suicide (Table 1). Although RCTs are considered the gold standard for establishing causation, they have significant limitations when studying suicide due to the rare occurrence of suicide events and the more likely exclusion of participants at higher suicide risk. Effective solutions are needed to include higher risk populations in clinical research focused on suicide prevention. The emergence of alternatives study designs such as target trial emulation and instrumental variable analysis may be particularly useful for suicide prevention research.^50, 51^ These study designs can emulate RCTs under appropriate conditions and may overcome some of the logistical challenges of conducting RCTs in high risk populations. However, limitations including potential for increased missing data and decreased data quality from lower reliability and validity compared to RCTs should be noted. Some studies have used these methods to investigate suicide outcomes in post-traumatic stress and depressive disorders, but there are no similar studies available for SCZ.^52-54^

We acknowledge some limitations in the current study. First, the rare occurrence of suicide, short-term trial durations, and exclusion of patients with higher suicide risk may confound the true occurrence of suicide events. Second, the inclusion of suicidal ideation as an outcome introduces heterogeneity risk given variability in symptom reporting. Third, the high heterogeneity across OLEs with some concerns of bias illustrate considerable differences across studies and limit the generalizability of single proportion meta-analysis results. Fourth, three studies could not be included due to the lack of data availability despite reaching out to authors. These studies focused on cariprazine (no trial registration number), lumateperone (NCT02469155), and oxaripiprazole (NCT01490086).^55, 56^ Additionally, although our search strategy was extensive and peer-reviewed, there is a possibility that other relevant studies may have been missed.

## 5. Conclusion

There is no current evidence to indicate that newer TGAs causally affect suicide outcomes for SCZ in either a positive or negative way. Subgroups analyses by antipsychotic agent, antipsychotic dose, and comparator arm showed no significant changes to suicide risk. While there was an increased incidence of primary and secondary suicide outcomes in OLEs of brexpiprazole and cariprazine, this signal should be interpreted with caution due to high heterogeneity and requires replication in other data. Our findings support the unique protective effect of clozapine against suicide and emphasize the critical need for more benign pharmacologic alternatives that are equally effective in preventing suicide in SCZ. Future studies into rare suicide outcomes should consider alternative high-quality study designs as well as study designs with larger sample sizes and longer term follow up. There is also a need to consider how to include high risk populations in clinical research focused on suicide prevention.

## Supporting information

PROSPERO Registration

Search Design

PRISMA Checklist

## Data Availability

All data produced in the present work are contained in the manuscript and supplementary materials

## 6. Data Availability

All data are available in the manuscript, figures, tables, and supplementary files.

## 7. Acknowledgements

None

## 8. Author Contributions

**Jeff W. Jin:** Methodology, Article Selection, Data Extraction, Risk of Bias Assessment, Statistical Analysis, Data Interpretation, Writing – Original Draft, Review & Editing, Visualization. **Charlotte J. Winkler:** Investigation, Data Extraction, Data Interpretation, Writing – Original Draft, Review & Editing. **Heather B. Blunt:** Methodology, Literature Search Design. **Natalie B. Riblet:** Conceptualization, Methodology, Investigation, Risk of Bias Assessment, Statistical Analysis, Data Interpretation, Writing – Original Draft, Review & Editing, Visualization, Supervision.

