## Supplementary Materials for "Suicide Risk of Third-Generation Antipsychotics in Persons with Schizophrenia and Schizoaffective Disorders: A Systematic Review and Meta-Analysis"

^3^ Mental Health, White River Junction VA Healthcare System, White River Junction, VT 05009, USA

^4^ Psychiatry, Geisel School of Medicine at Dartmouth College, Hanover, NH 03755, USA

^5^ Dartmouth Institute, Geisel School of Medicine at Dartmouth College, Hanover, NH 03755, USA

**Inclusion Criteria**

1. Human Subjects
2. English Language
3. Randomized Controlled Trial or Open Label Extension Trial Design
4. Diagnostic and Statistical Manual of Mental Disorders Fourth Edition (DSM-IV) diagnosis of schizophrenia, schizoaffective disorder, schizophreniform disorder, unspecified psychosis
5. Third generation antipsychotic intervention (oxaripiprazole, brexpiprazole, cariprazine, lumateperone)
6. Reported data on suicide outcomes (deaths, attempts, and ideation)

**Exclusion Criteria**

1. Cannabis or substance-induced psychosis only diagnoses were excluded
2. Prodromal psychosis states including early phase psychosis, sub-clinical psychosis, and clinical high-risk for psychosis were excluded
3. Self-reported psychosis diagnoses were excluded
4. Studies focused on aripiprazole were excluded
5. Data on non-suicidal self-injurious behavior were excluded from analysis

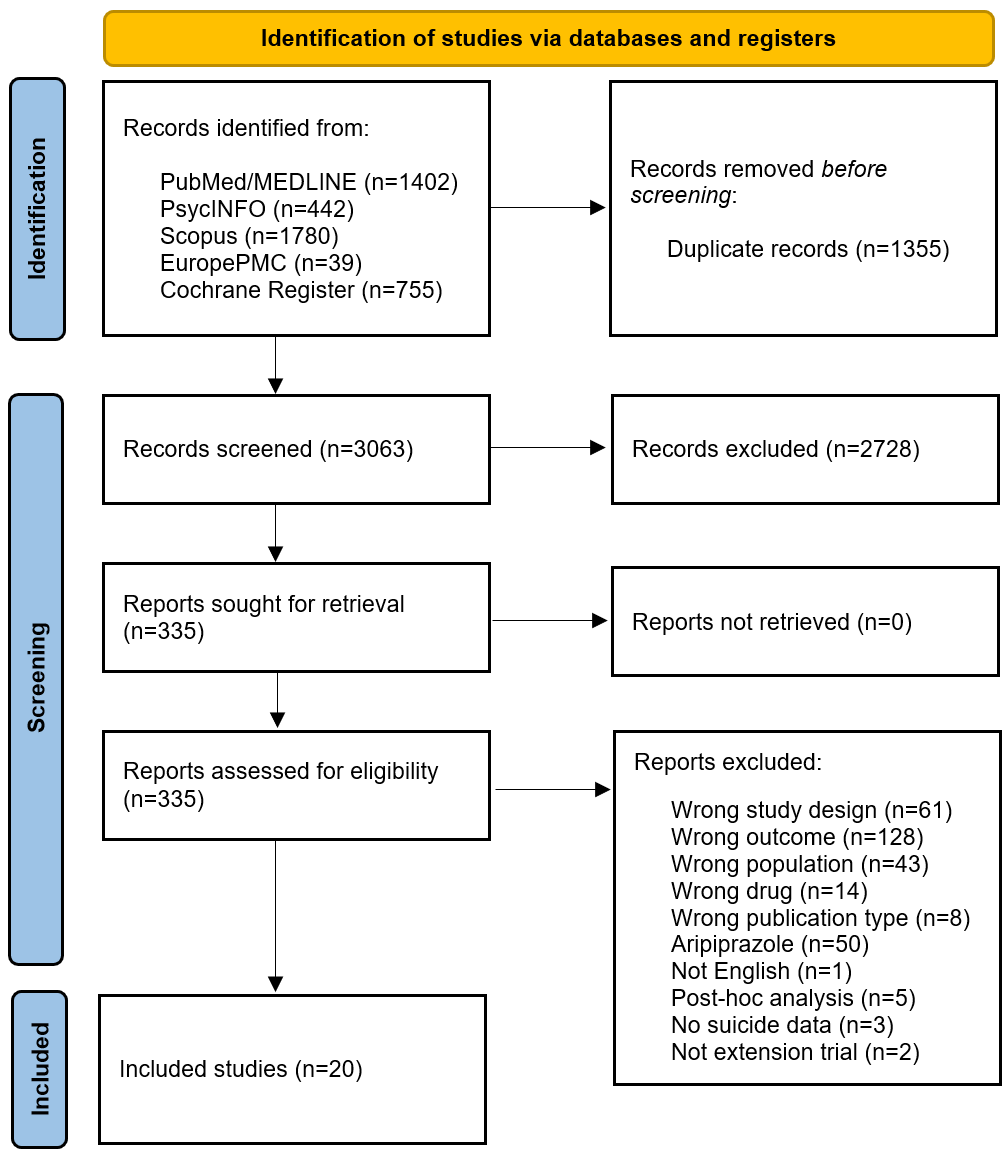
**Supplementary Figure S1.** PRISMA Flow Diagram for study inclusion.

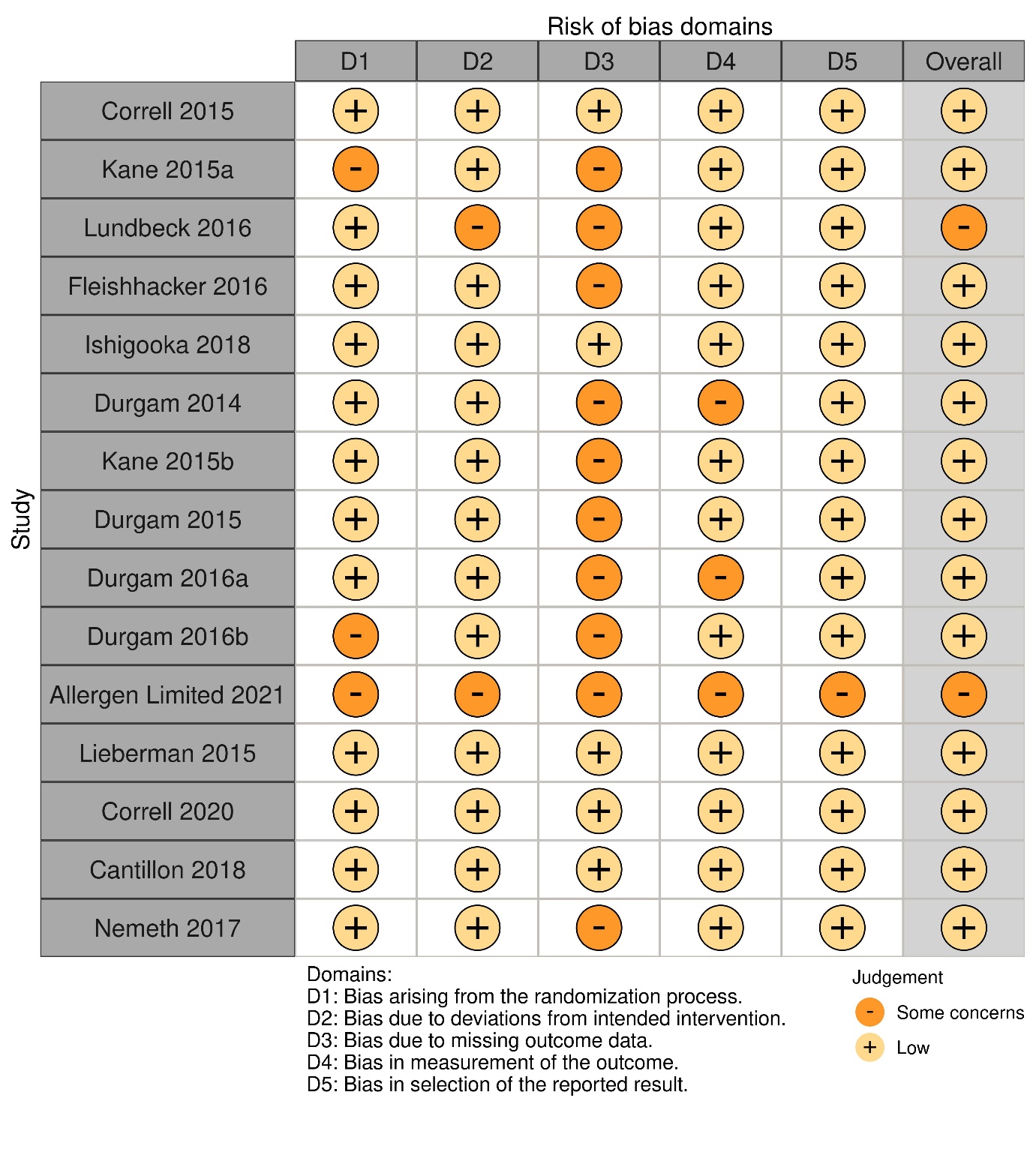
**Supplementary Table S1.** Risk of Bias Assessment for Randomized Controlled Trials by Domain

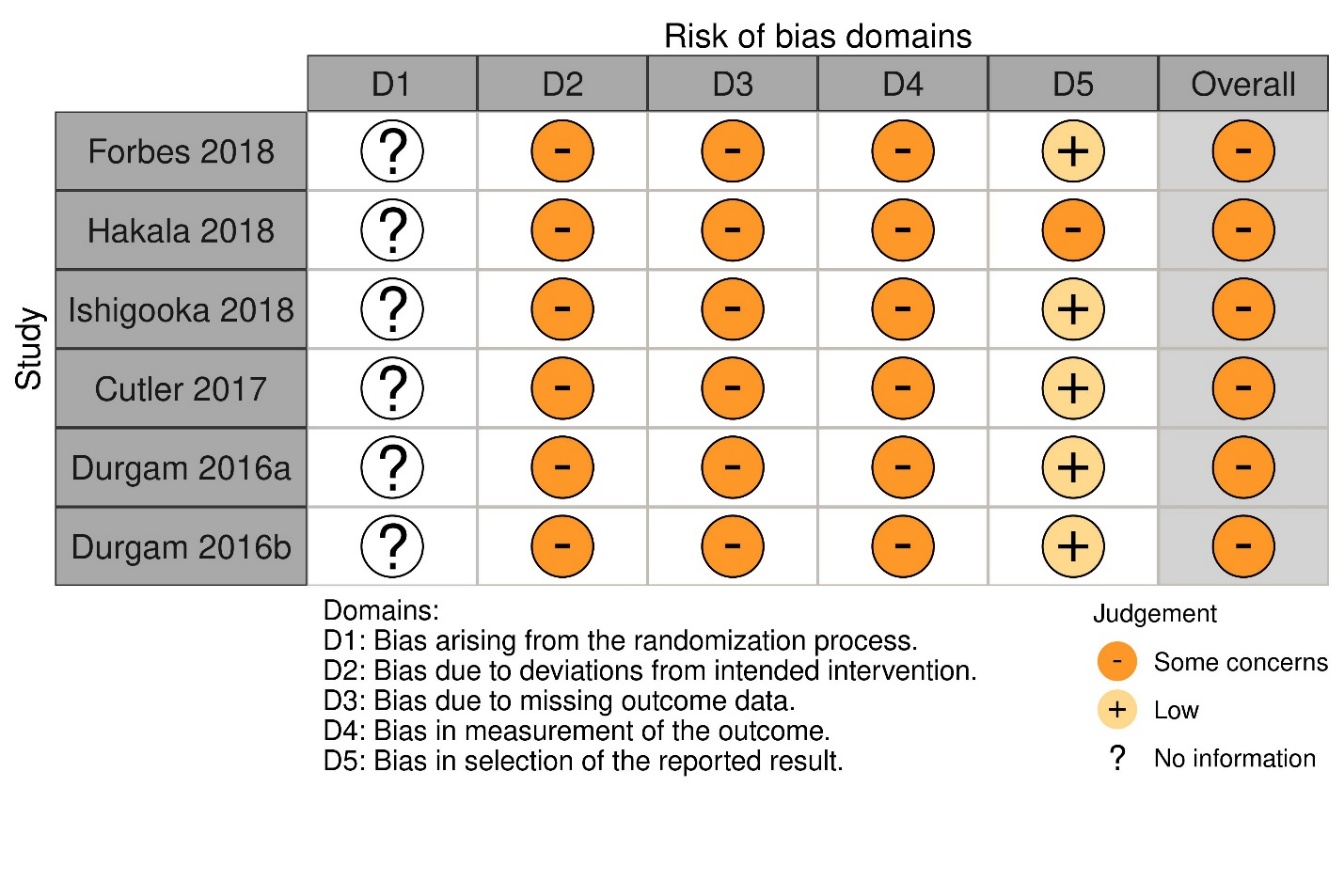
**Supplementary Table S2.** Risk of Bias Assessment for Open Label Extension Trials by Domain

*****No information for D1 due to lack of randomization in open label trials

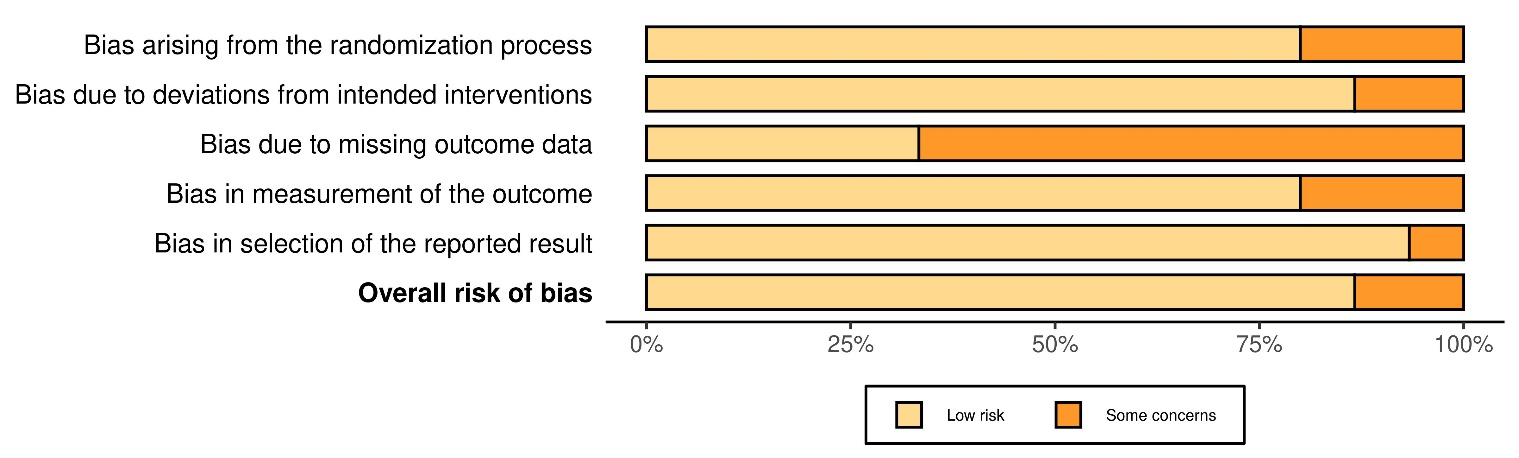
**Supplementary Figure S2.** Risk of Bias of Included Randomized Controlled Trials.

**Supplementary Figure S3.** Risk of Bias of Included Open-Label Extensions Trials.

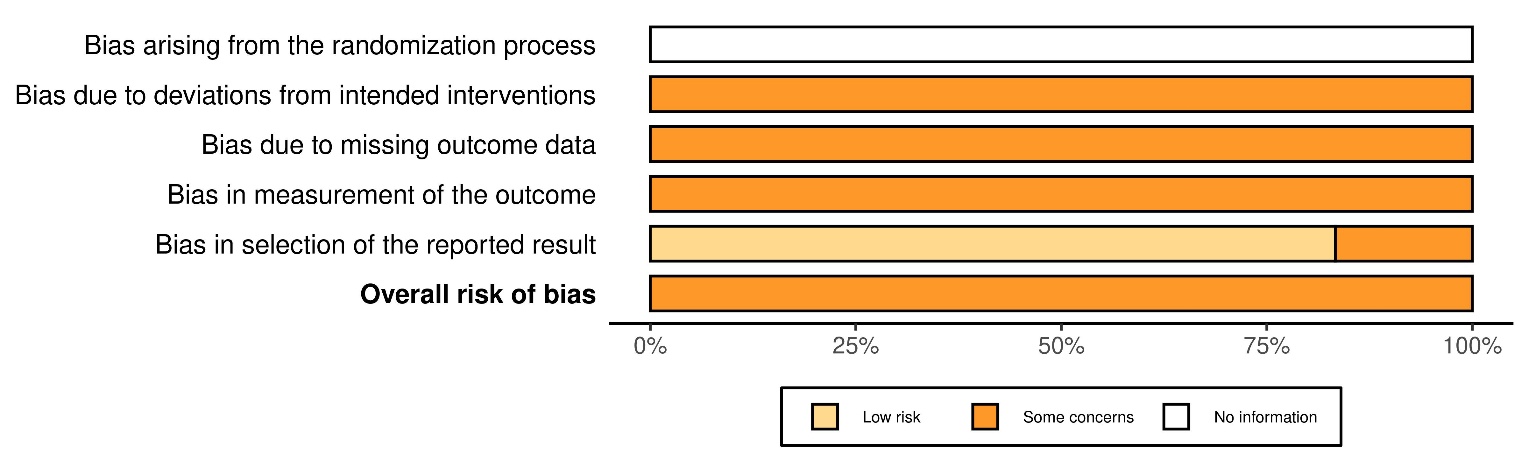

*****No information for randomization bias due to lack of randomization in open label trials

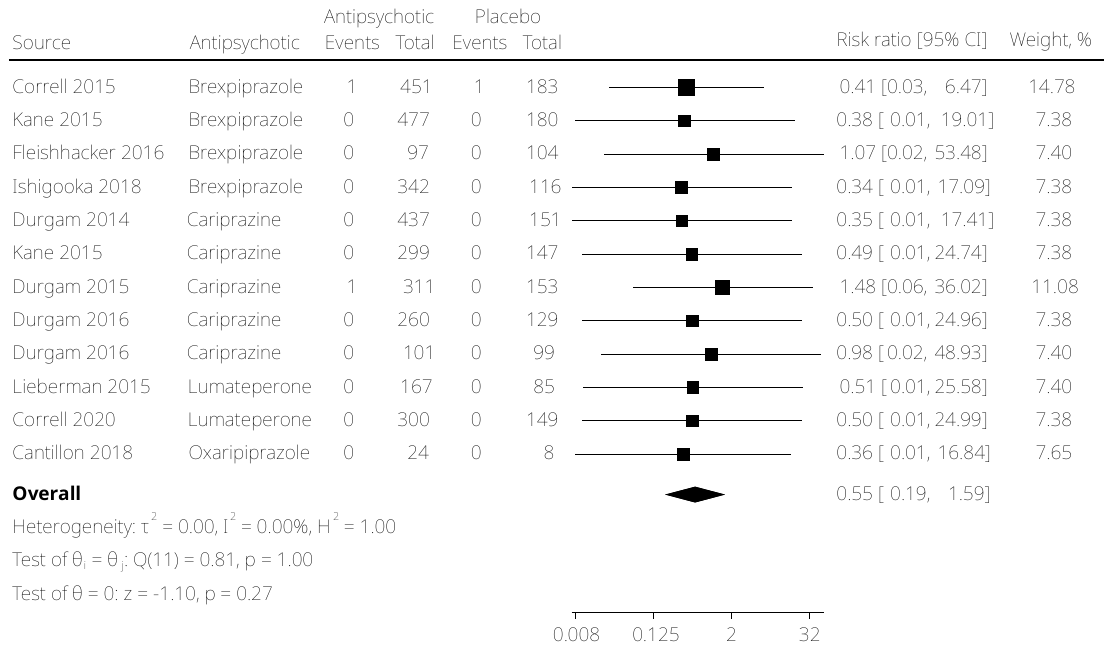
**Supplementary Figure S4.** Sensitivity Analysis of Suicide Deaths and Attempts in Randomized Controlled Trials

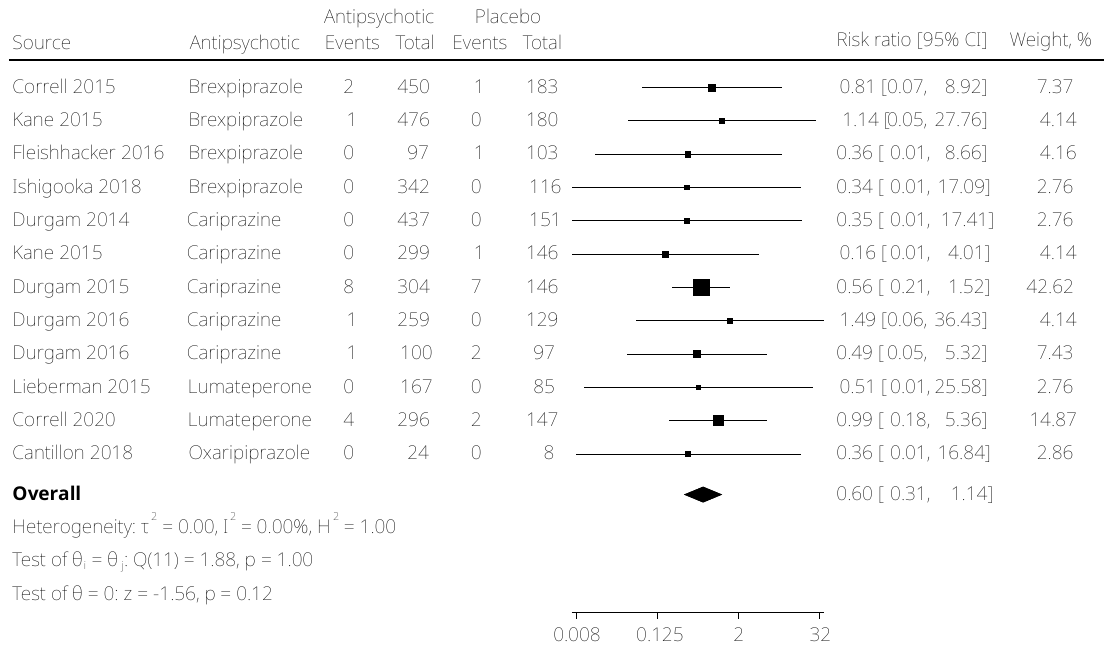
**Supplementary Figure S5.** Sensitivity Analysis of Suicide Deaths, Attempts, and Ideation in Randomized Controlled Trials

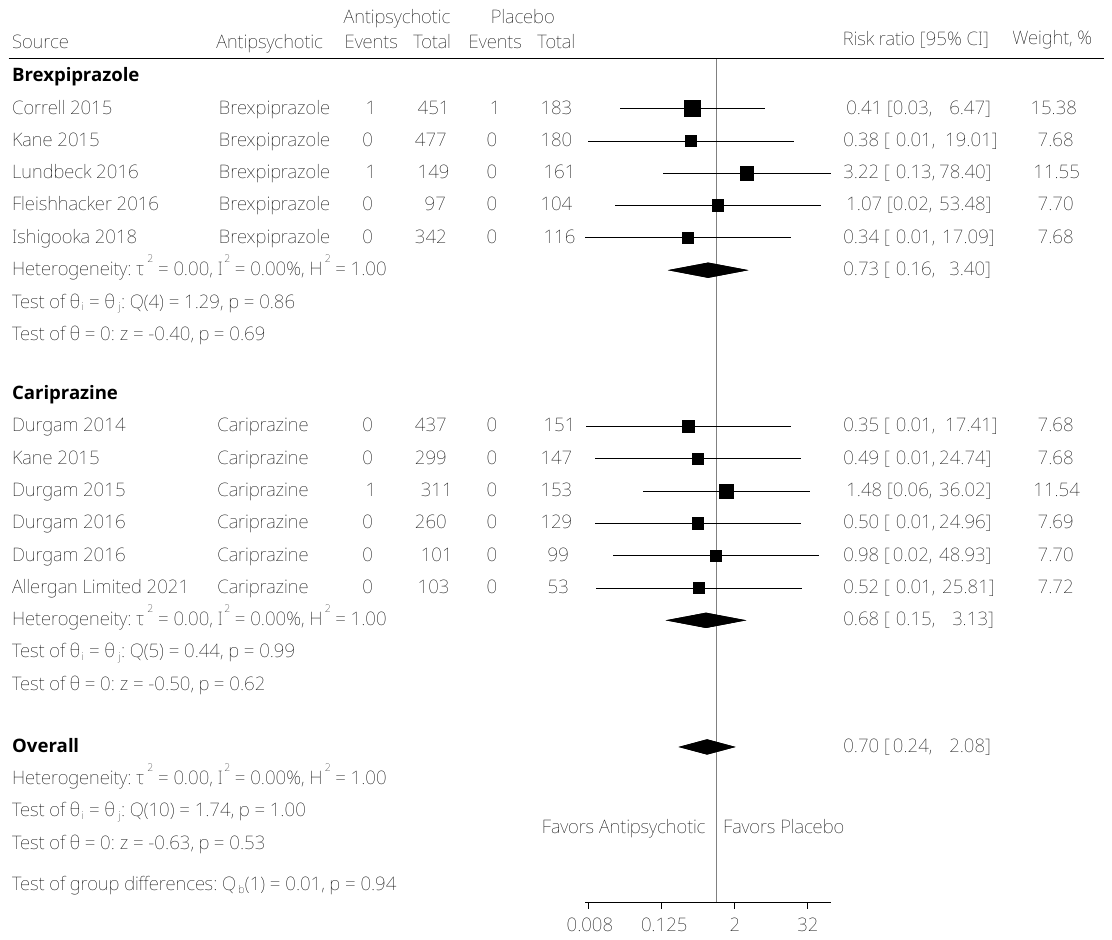
**Supplementary Figure S6.** Subgroup Analysis of Suicide Deaths and Attempts in Randomized Controlled Trials by Antipsychotic

**Supplementary Figure S7.** Sensitivity Analysis of Subgroup Analysis of Suicide Deaths and Attempts in Randomized Controlled Trials by Antipsychotic

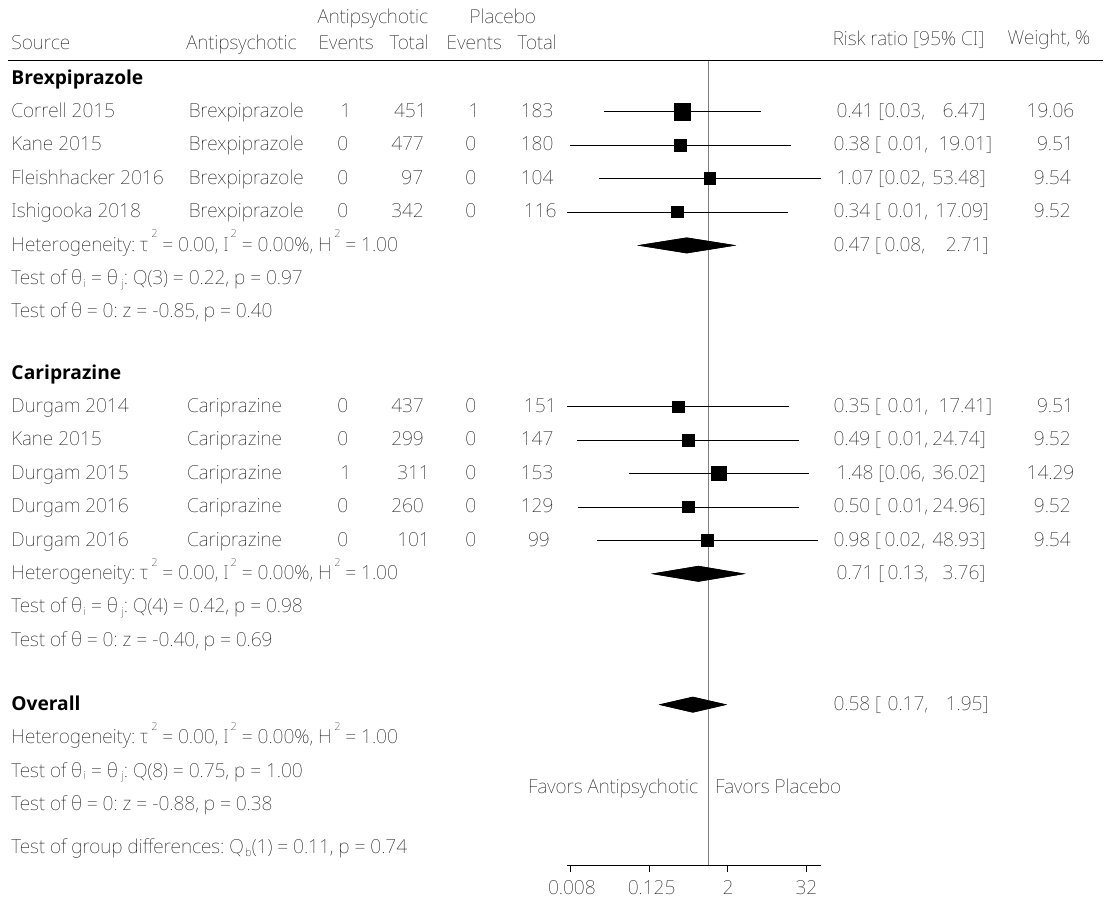

**Supplementary Figure S8.** Subgroup Analysis of Suicide Deaths, Attempts, and Ideation in Randomized Controlled Trials by Antipsychotic

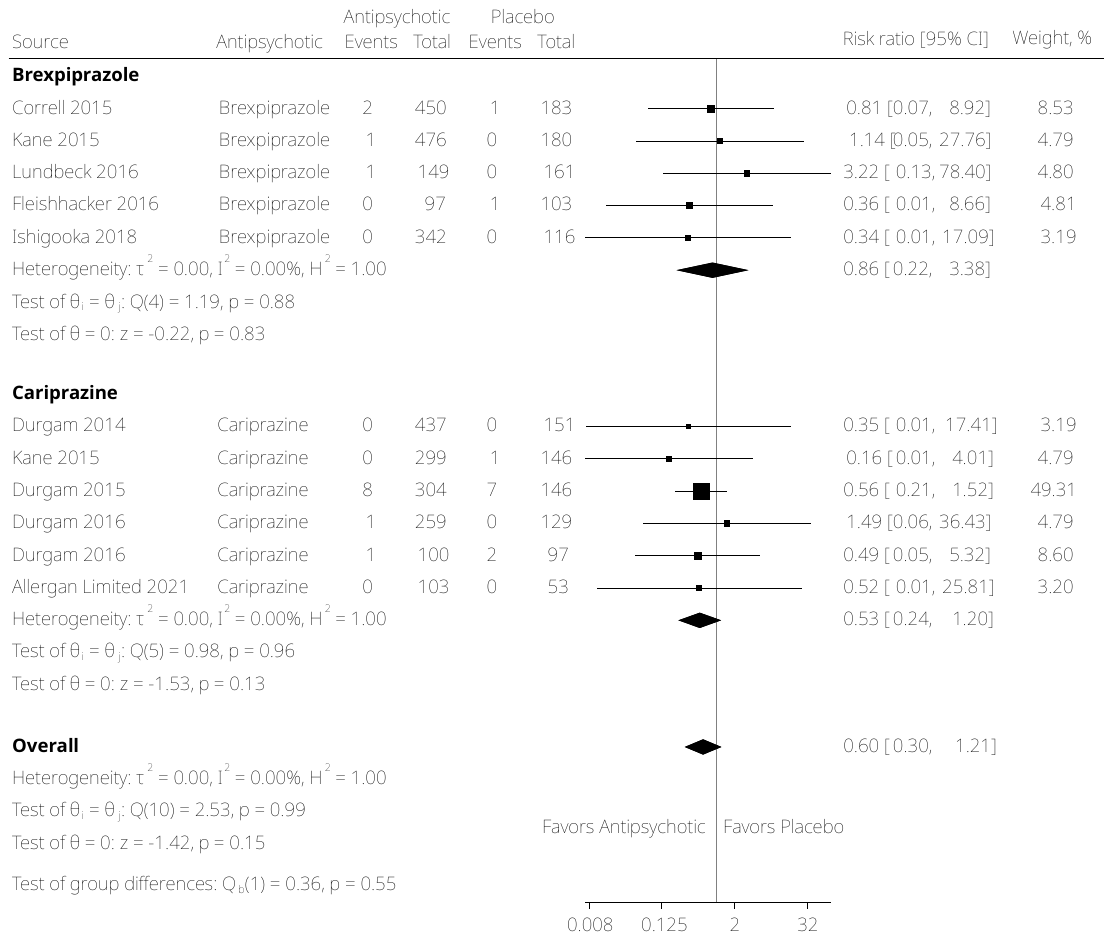

**Supplementary Figure S9.** Sensitivity Analysis of Subgroup Analysis of Suicide Deaths, Attempts, and Ideation in Randomized Controlled Trials by Antipsychotic

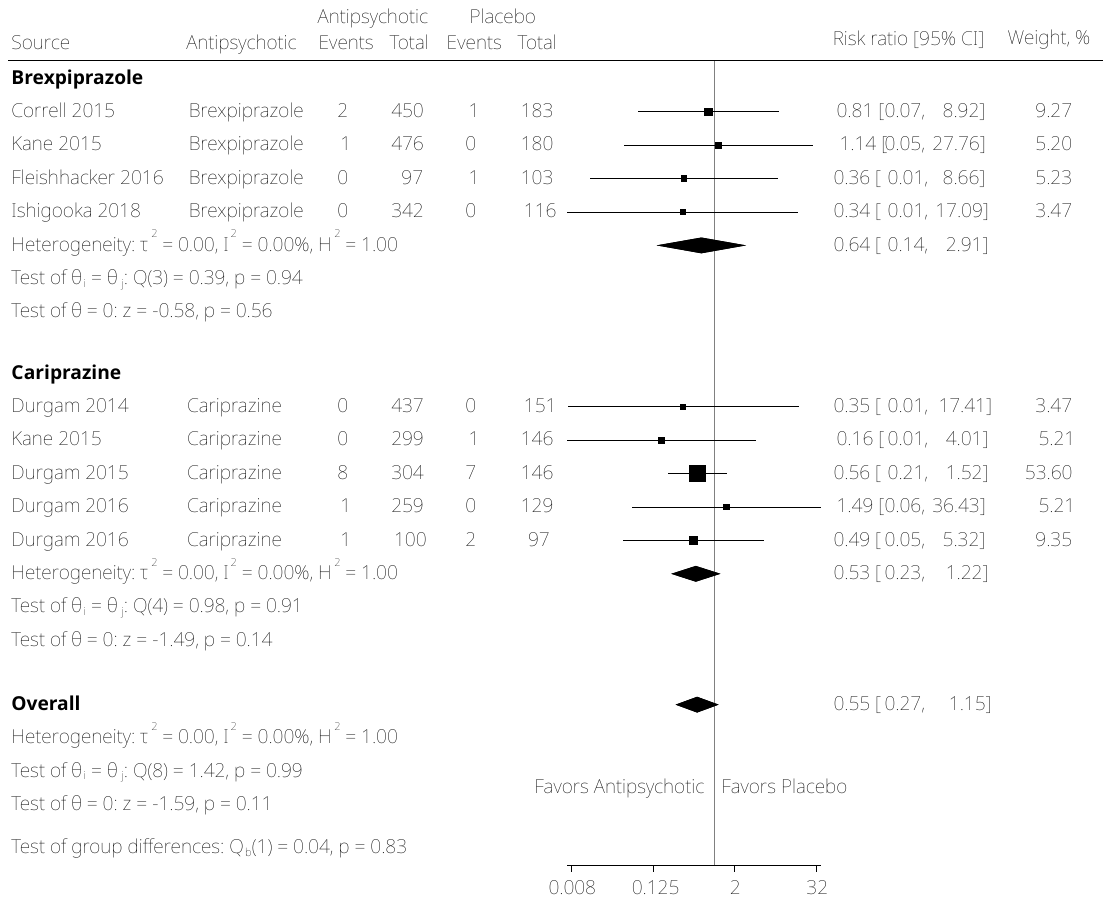

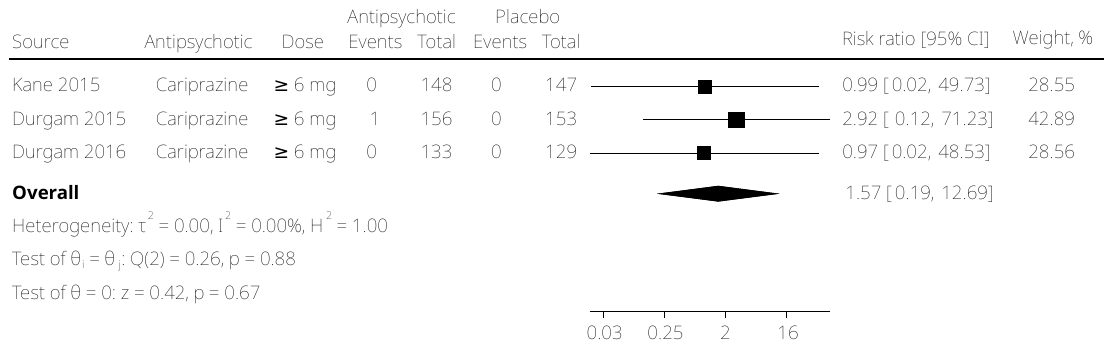

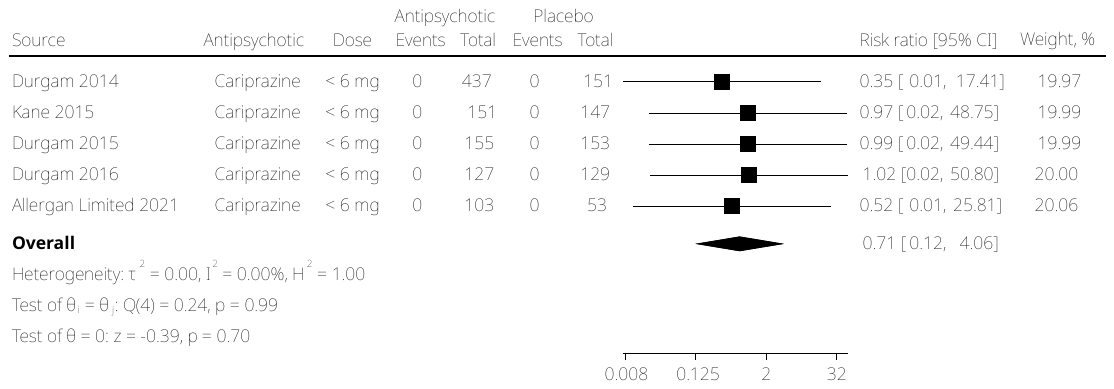

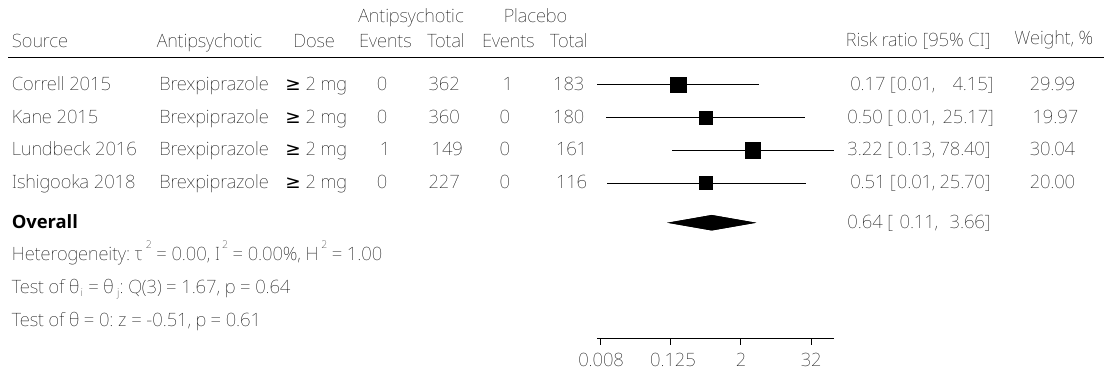

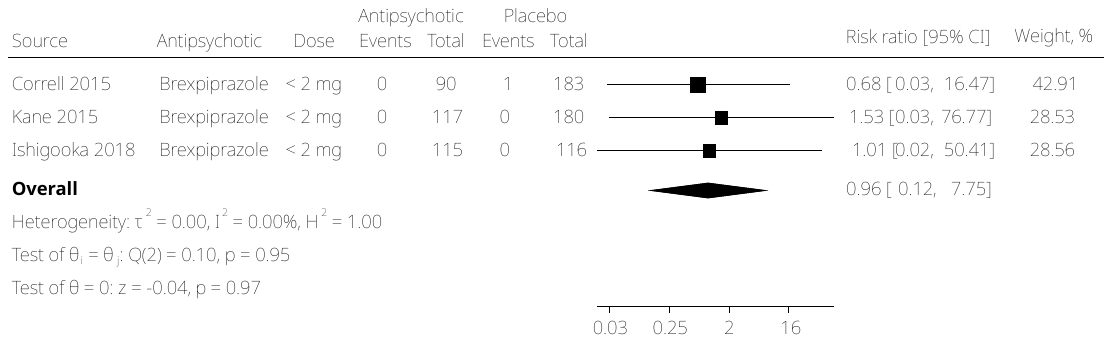
**Supplementary Figure S10.** Subgroup Analysis of Suicide Deaths and Attempts in Randomized Controlled Trials by Antipsychotic Dose

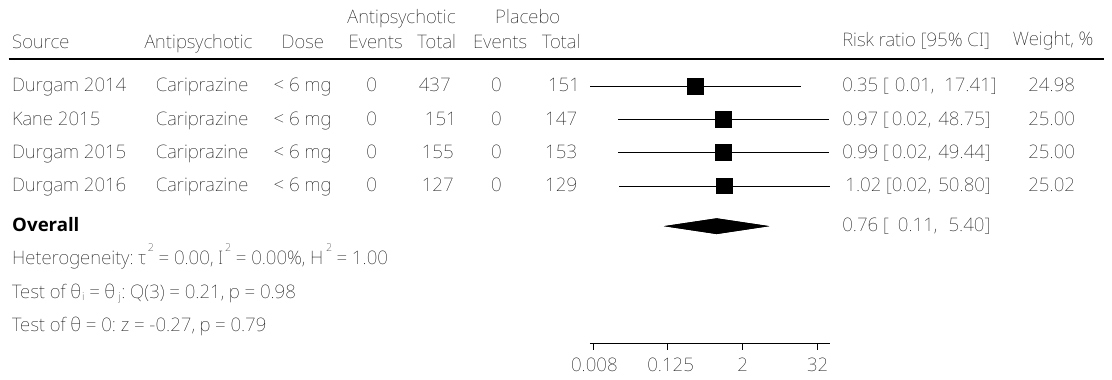

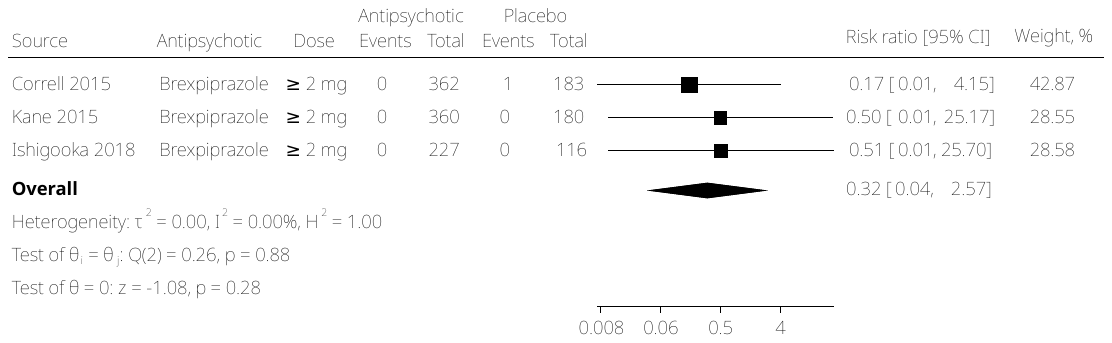
**Supplementary Figure S11.** Sensitivity Analysis of Subgroup Analysis of Suicide Deaths and Attempts in Randomized Controlled Trials by Antipsychotic Dose

*****No sensitivity analyses needed for Brexpiprazole < 2 mg and Cariprazine ≥ 6 mg due to lack of studies with “some concerns” or “high risk” of bias

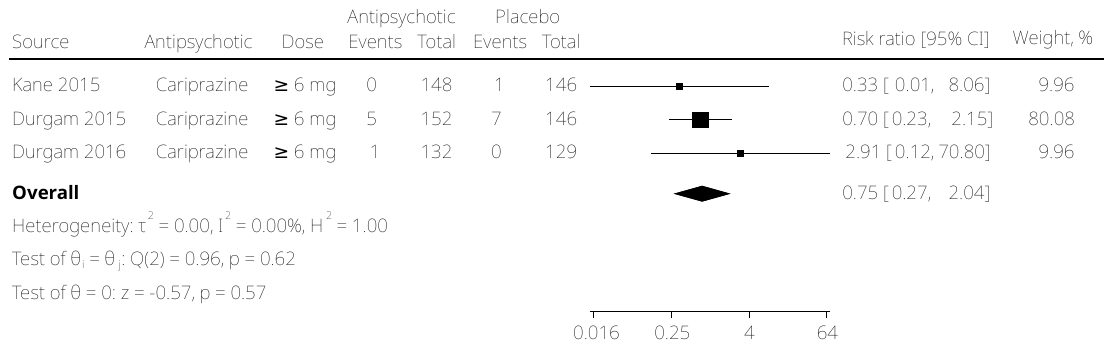

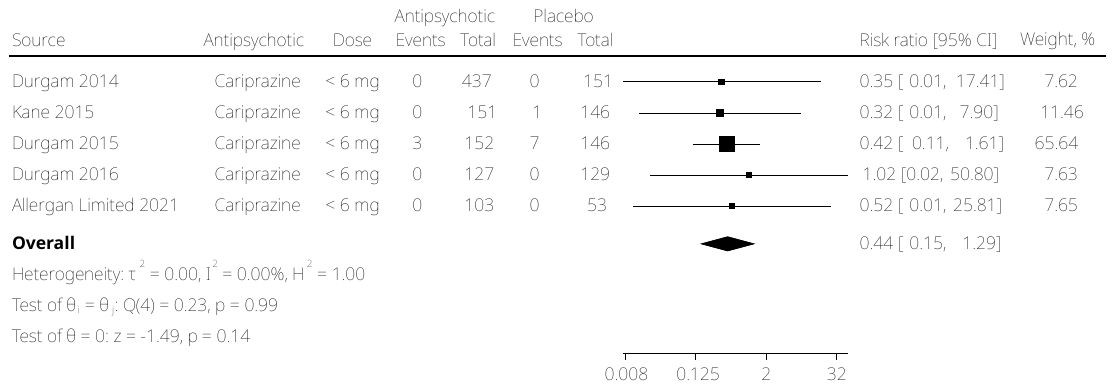

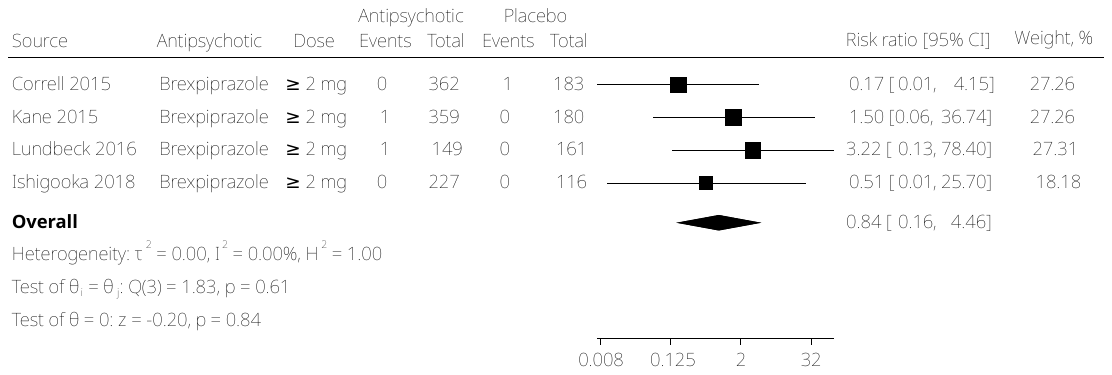
**Supplementary Figure S12.** Subgroup Analysis of Suicide Deaths, Attempts, and Ideation in Randomized Controlled Trials by Antipsychotic Dose

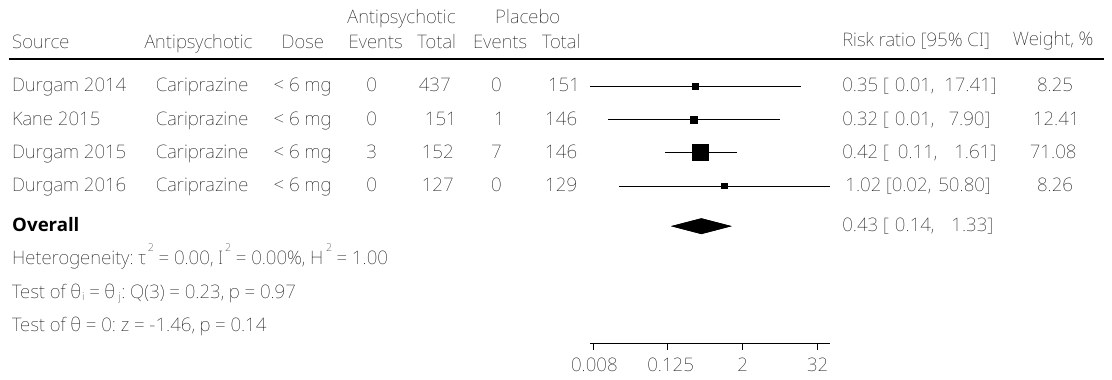
**Supplementary Figure S13.** Sensitivity Analysis of Subgroup Analysis of Suicide Deaths, Attempts, and Ideation in Randomized Controlled Trials by Antipsychotic Dose

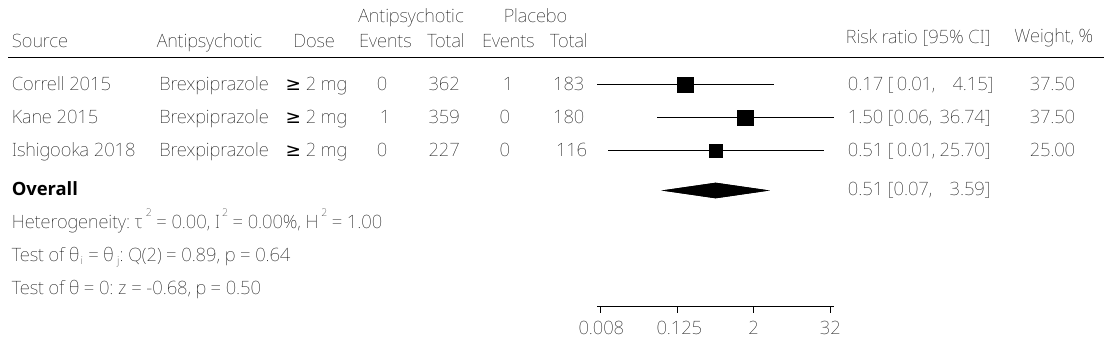
*****No sensitivity analyses needed for Brexpiprazole < 2 mg and Cariprazine ≥ 6 mg due to lack of studies with “some concerns” or “high risk” of bias

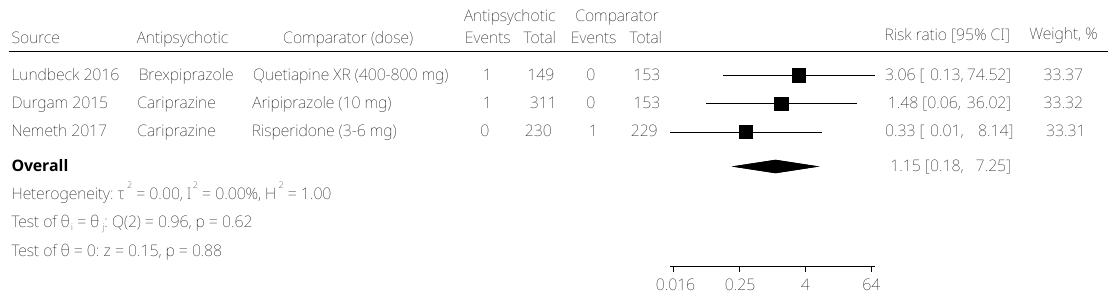
**Supplementary Figure S14.** Subgroup Analysis of Suicide Deaths and Attempts in Randomized Controlled Trials by Comparator

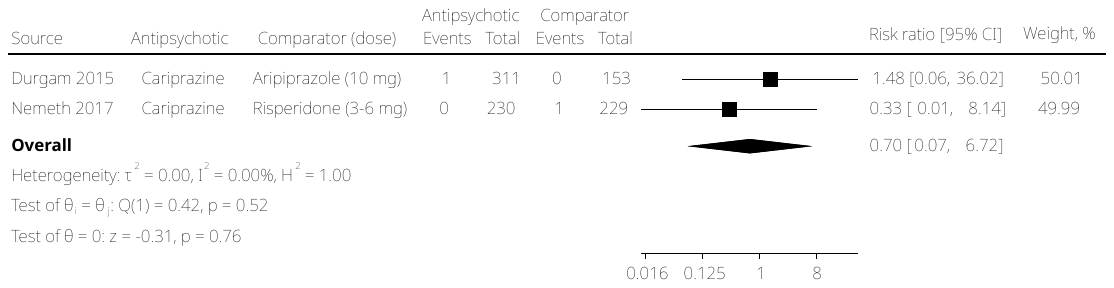
**Supplementary Figure S15.** Sensitivity Analysis of Subgroup Analysis of Suicide Deaths and Attempts in Randomized Controlled Trials by Comparator

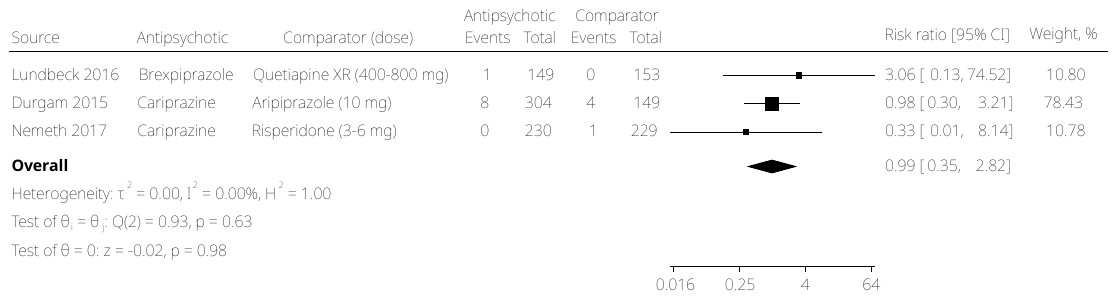
**Supplementary Figure S16.** Subgroup Analysis of Suicide Deaths, Attempts, and Ideation in Randomized Controlled Trials by Comparator

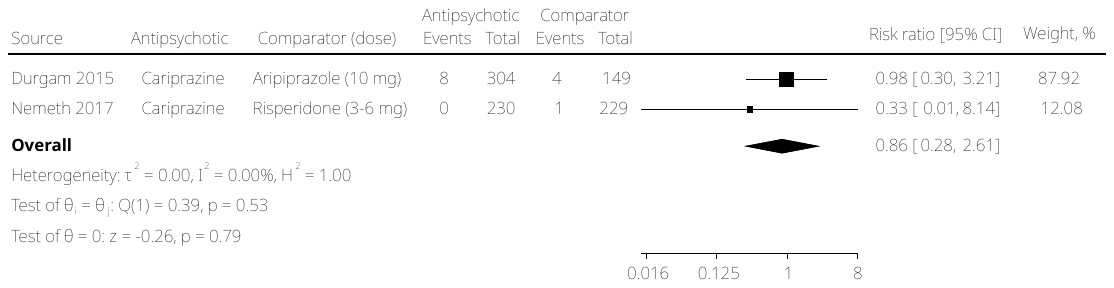
**Supplementary Figure S17.** Sensitivity Analysis of Subgroup Analysis of Suicide Deaths, Attempts, and Ideation in Randomized Controlled Trials by Comparator

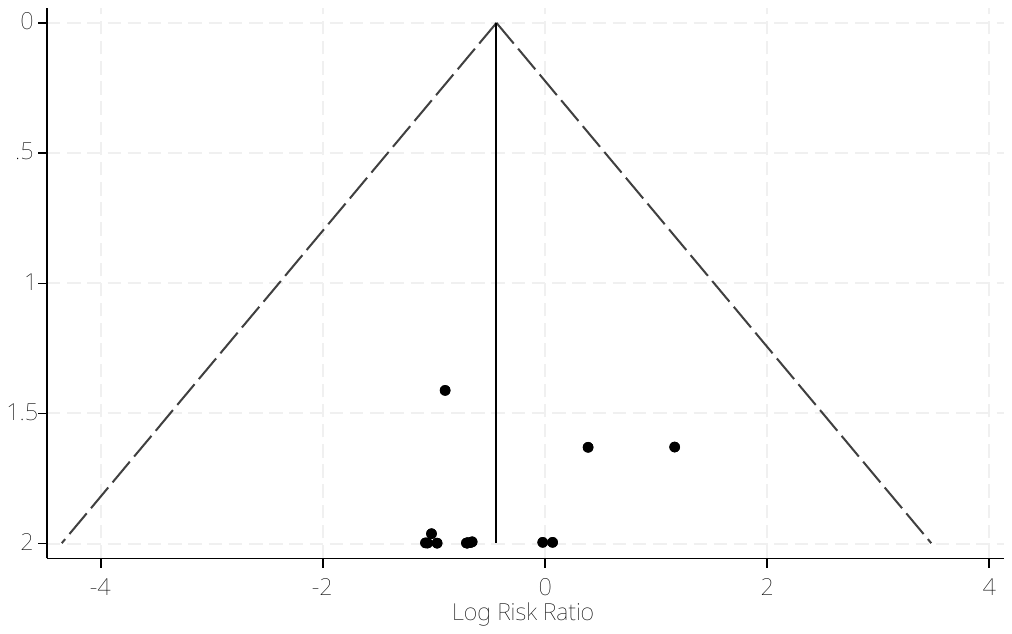
**Supplementary Figure S18.** Overall Funnel Plot for Included Randomized Controlled Trials

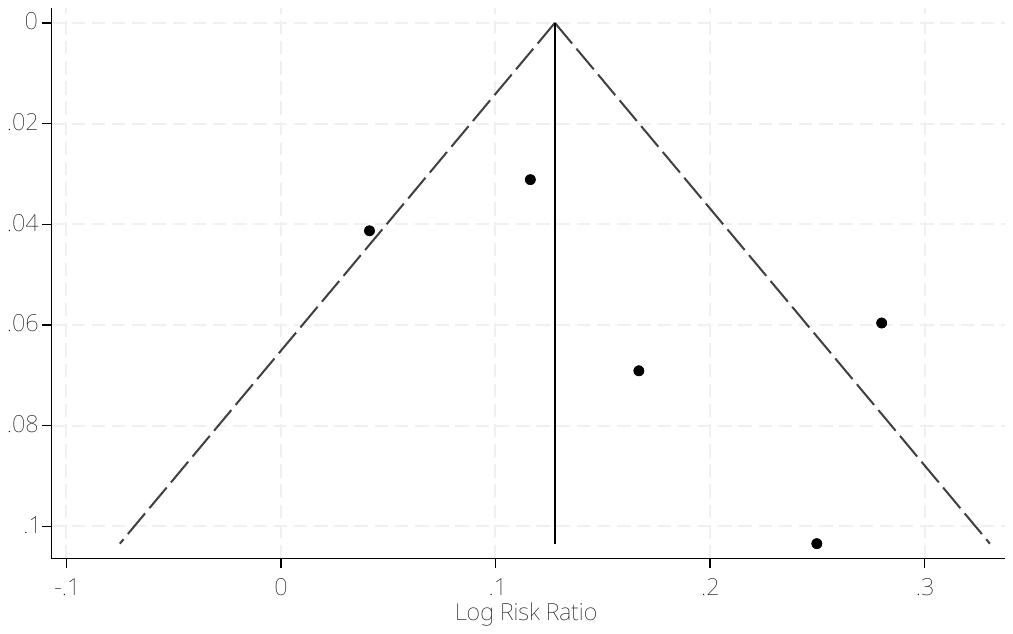
**Supplementary Figure S19.** Overall Funnel Plot for Included Open Label Extension Trials
