## Supplementary material for "Suicide Risk of Third-Generation Antipsychotics in Persons with Schizophrenia and Schizoaffective Disorders: A Systematic Review and Meta-Analysis": PROSPERO Registration

NIHR | National Institute for Health and Care Research

PROSPERO
International prospective register of systematic reviews

### Effect of Third Generation Antipsychotics on Suicide Risk in Persons with Schizophrenia Spectrum Disorders

*Jeff Jin, Charlotte Winkler, Heather Blunt, Natalie Riblet*

Review methods were amended after registration. Please see the revision notes and previous versions for detail.
To enable PROSPERO to focus on COVID-19 submissions, this registration record has undergone basic automated checks for eligibility and is published exactly as submitted. PROSPERO has never provided peer review, and usual checking by the PROSPERO team does not endorse content. Therefore, automatically published records should be treated as any other PROSPERO registration. Further detail is provided here.

**Citation** 1 change
Jeff Jin, Charlotte Winkler, Heather Blunt, Natalie Riblet. Effect of Third Generation Antipsychotics on Suicide Risk in Persons with Schizophrenia Spectrum Disorders. PROSPERO 2023 CRD42023449336. Available from https://www.crd.york.ac.uk/PROSPERO/view/CRD42023449336.

#### REVIEW TITLE AND BASIC DETAILS

**Review title** 1 change
Effect of Third Generation Antipsychotics on Suicide Risk in Persons with Schizophrenia Spectrum Disorders

**Review objectives** 1 change
In persons with schizophrenia and schizoaffective disorder, what effects do newer third generation antipsychotics have on suicidal risk?

- P - Schizophrenia Spectrum Illness
- I - 3rd generation antipsychotics (cariprazine, lumateperone, brexpiprazole, oxaripiprazole)
- C - Placebo vs. other antipsychotics
- O - Suicide attempts and completions

**Keywords**
Antipsychotics; Meta-analysis; Psychotic disorders; Suicide; Schizophrenia; Schizoaffective

#### SEARCHING AND SCREENING

##### Searches 1 change

- Databases: PubMed/MEDLINE, PsycINFO (EBSCO), Cochrane Central Register of Controlled Trials (Wiley), Scopus, Europe PMC (preprints), ClinicalTrials.gov
- Search Dates: Inception to December 1st, 2023

##### Study design 1 change

Both randomized and nonrandomized study types will be included.

*Included*
Randomized Controlled Trials, Open Label Extension Trials

*Excluded*
The following studies will be excluded:

- Non-Human subject studies
- Non-English publications
- Wrong study design (not randomized controlled trial or open label extension trial)
- Wrong diagnosis (not schizophrenia or schizoaffective disorder)
- Wrong intervention (not third-generation antipsychotic)
- Studies focused on aripiprazole
- Studies that do not report data on suicide outcomes (deaths, attempts, ideation)

##### Link to search strategy

A full search strategy has been uploaded to PROSPERO. The PDF may be accessed through this link https://www.crd.york.ac.uk/PROSPEROFILES/91ac3f42b6ec6baf29175eed299eef04.pdf.

#### ELIGIBILITY CRITERIA

##### Condition or domain being studied 1 change

*Schizophrenia; Schizoaffective Disorder; Schizophreniform Disorder; Schizophrenic Related Disorders; Suicide; Suicidal Behavior; Suicidal Ideation*

- Schizophrenia spectrum illness as defined by the National Institute of Mental Health and Diagnostic and Statistical Manual of Mental Disorders, fifth edition (DSM-V) as diagnoses of schizophrenia, schizoaffective disorder, or an unspecified schizophrenia spectrum/other psychotic disorder.
- Schizophrenia - a mental disorder characterized by disruptions in thought processes, perceptions, emotional responsiveness, and social interactions
- Schizoaffective disorder - a chronic mental health condition characterized primarily by symptoms of schizophrenia, such as hallucinations or delusions, and symptoms of a mood disorder, such as mania and depression.
- Unspecified schizophrenia spectrum/other psychotic disorder - a mental health condition characterized by symptoms of a schizophrenia spectrum and other psychotic disorder that

cause clinically significant distress or impairment in social, occupational, or other important areas of functioning predominate but do not meet the full criteria for any of the disorders in the schizophrenia spectrum and other psychotic disorders class

##### Population 1 change

*Included*

- Inclusion: Patients with schizophrenia and related psychotic spectrum illnesses (as diagnosed by any recognized diagnostic criteria for schizophrenia, schizoaffective disorder, schizophreniform, or an unspecified schizophrenia spectrum/other psychotic disorder)
- Exclusion: Non-schizophrenia spectrum illness population

##### Intervention(s) or exposure(s) 1 change

*Included*

- Third generation antipsychotics - a class of psychotropic medication used to treat psychosis that is characterized by a common pharmacodynamic feature of partial dopamine D2 agonism
- Current medications of this class include oxaripiprazole, brexpiprazole, lumateperone, and cariprazine

*Excluded*
Aripiprazole has been excluded to focus the meta-analysis on newer agents

##### Comparator(s) or control(s) 1 change

*Included*

- Placebo
- Non-third generation antipsychotics (treatment as usual)

##### Context 1 change
Studies in hospital, partial hospitalization, inpatient, and outpatient psychiatric care settings will be included.

#### OUTCOMES TO BE ANALYSED

##### Main outcomes 1 change

- Primary Outcome: suicide deaths and attempts
- Secondary Outcome: suicide deaths, attempts, and ideation
- Instruments: Validated measurements to assess suicide risk (such as the Columbia Suicide Severity Rating Scale)
- Time points: Study dependent
- Effect measure: Risk Ratio

##### Additional outcomes 1 change
Not applicable

#### DATA COLLECTION PROCESS

##### Data extraction (selection and coding) 1 change

- Study selection: Two reviewers will apply the eligibility criteria by independently screening records for inclusion and cross-checking selection results after screening completion. Both researchers will be blinded to the other's decisions during the selection process. Disagreements will be formally discussed and resolved by consensus. In the case a disagreement cannot be resolved, a clinically-informed third party will be consulted for resolution by majority. Rayyan QCRI is the current preferred mechanism for screening decision records.
- Data extraction: Information about study design, participant demographics, intervention type, suicide outcome, and study results will be extracted by comprehensive and in-depth article review after the screening process. Two researchers will be extracting the data while a third will be reviewing the extracted data for accuracy. Disagreements will be formally discussed and resolved by consensus. In the case a disagreement cannot be resolved, a clinically-informed third party will be consulted for resolution by majority. Missing data will be re-visited based on appearance. Data will be recorded in a shared protected-spreadsheet.

##### Risk of bias (quality) assessment 1 change

Two review authors will independently assess the risk of bias of included studies by using the considering the followingcharacteristics, as recommended by the International Cochrane Collaboration:

- Randomization sequence generation
- Treatment allocation concealment
- Blinding
- Completeness of outcome data
- Selective outcome reporting
- Other sources of bias

The revised Cochrane Risk-Of-Bias tool for randomized trials (RoB2) will be used. Disagreements will be formally discussed and resolved by consensus. In the case a disagreement cannot be resolved, a clinically-informed third party will be consulted for resolution by majority. The level of risk of bias in each of these domains will be presented separately for each study in tables in the final review publication.

#### PLANNED DATA SYNTHESIS

##### Strategy for data synthesis 1 change

- Software: Stata SE18
- Statistical Significance: p <0.05
- Effects Size Estimates: Risk Ratios, 95% Confidence Intervals
- For randomized controlled trials, a random-effects meta-analysis will be used

- For open label extension studies, a single proportion meta-analysis will be used
- For study arms with zero events, a continuity correction of 0.5 was added to prevent mathematical impossibility (standard in Stata SE18)
- Study heterogeneity will be explored statistically (defined as $I$-squared > 50% or $p < 0.1$)
- Publication bias will be assessed with funnel plots and Egger's tests

##### Analysis of subgroups or subsets 1 change

- Sensitivity analyses will be conducted to exclude randomized controlled trials with "some concerns" or "high risk" of bias
- Subgroup analyses will be conducted for randomized controlled trials to explore the impact of dose, drug type, and choice of comparator arm on suicide outcomes

#### REVIEW AFFILIATION, FUNDING AND PEER REVIEW

##### Review team members

**Dr Jeff Jin** (review guarantor and contact) Dartmouth-Hitchcock Medical Center. United States of America.

No conflict of interest declared.

**Dr Charlotte Winkler**. Dartmouth-Hitchcock Medical Center. United States of America.

No conflict of interest declared.

**Mrs Heather Blunt**. Dartmouth-Hitchcock Medical Center. United States of America.

No conflict of interest declared.

**Dr Natalie Riblet**. Dartmouth-Hitchcock Medical Center. United States of America.

No conflict of interest declared.

##### Named contact

**Dr Jeff Jin**. Dartmouth-Hitchcock Medical Center. United States of America.

##### Review affiliation

Dartmouth-Hitchcock Medical Center

##### Funding source

Not applicable

#### TIMELINE OF THE REVIEW

##### Review timeline

Start date: 14 August 2023. End date: 30 June 2026.

##### Date of first submission to PROSPERO

09 August 2023

#### Date of registration in PROSPERO

21 August 2023

### CURRENT REVIEW STAGE

#### Publication of review results

The intention is to publish the review once completed.The review will be published in English

#### Stage of the review at this submission 1 change

| Review stage | Started | Completed |
|---|---|---|
| Pilot work | ✓ | ✓ |
| Formal searching/study identification | ✓ | ✓ |
| Screening search results against inclusion criteria | ✓ | ✓ |
| Data extraction or receipt of IPD | ✓ | ✓ |
| Risk of bias/quality assessment | ✓ | ✓ |
| Data synthesis | ✓ | ✓ |

#### Review status

The review is completed.

### ADDITIONAL INFORMATION

#### PROSPERO version history 1 change

- Version 3.0, published 04 Feb 2026
- Version 2.0, published 20 Nov 2025
- Version 1.4, published 20 Jan 2025
- Version 1.3, published 12 Feb 2024
- Version 1.2, published 05 Dec 2023
- Version 1.1, published 21 Aug 2023
- Version 1.0, published 21 Aug 2023

#### Review conflict of interest

None known

#### Country

United States of America

#### Medical Subject Headings

Antipsychotic Agents; Humans; Schizophrenia; Suicidal Ideation; Suicide, Attempted; brexpiprazole; cariprazine; lumateperone; Suicide, Completed

#### Revision note 1 change

Revised to update the current status of the review and to provide additional detail to the methods section for further clarification and detail.

##### Disclaimer

The content of this record displays the information provided by the review team. PROSPERO does not peer review registration records or endorse their content.

PROSPERO accepts and posts the information provided in good faith; responsibility for record content rests with the review team. The guarantor for this record has affirmed that the information provided is truthful and that they understand that deliberate provision of inaccurate information may be construed as scientific misconduct.

PROSPERO does not accept any liability for the content provided in this record or for its use. Readers use the information provided in this record at their own risk.

Any enquiries about the record should be referred to the named review contact
