## Supplementary material for "Suicide Risk of Third-Generation Antipsychotics in Persons with Schizophrenia and Schizoaffective Disorders: A Systematic Review and Meta-Analysis": Search Design

### Search Strategies

#### Databases

| Database | Platform | Years covered | Date conducted | # results |
|---|---|---|---|---|
| Medline | PubMed | 1946 - current | December 1, 2023 | 1402 |
| APA PsycInfo | EBSCO | 1806-current | December 1, 2023 | 442 |
| Cochrane Central Register of Controlled Trials | Wiley | Issue 11, Nov 2023 | December 1, 2023 | 755 |
| EuropePMC | EuropePMC.org | 2018-current | December 1, 2023 | 39 |
| Scopus | Elsevier | 1788-current | December 1, 2023 | 1780 |
| Total | | | | 4418 |
| Total with duplicates removed | | | | **3063** |

#### Search Strategies

All searches performed on December 1, 2023
Searched by Heather Blunt, Dartmouth Biomedical Libraries
Peer reviewed by Elaina Vitale, Dartmouth Biomedical Libraries

#### PubMed

| Search | Query | Results |
|---|---|---|
| #5 | Search: #1 AND #2 AND #3 AND #4 | 1,402 |
| #4 | Search: ((randomized controlled trial[pt] OR controlled clinical trial[pt] OR randomized[tiab] OR randomised[tiab] OR placebo[tiab] OR drug therapy[sh] OR randomly[tiab] OR trial[tiab] OR groups[tiab]) NOT (animals[mh] NOT humans[mh])) | 5,179,129 |
| #3 | Search: "Schizophrenia Spectrum and Other Psychotic Disorders"[Mesh] OR "Schizoid Personality Disorder"[Mesh] OR "Schizotypal Personality Disorder"[Mesh] OR schizo*[tiab] OR Psychotic[tiab] OR Psychosis[tiab] OR Psychoses[tiab] OR deficit syndrome[tiab] | 251,807 |

| | | |
|---|---|---|
| #2 | Search: "Aripiprazole"[Mesh] OR "brexpiprazole" [Supplementary Concept] OR "cariprazine" [Supplementary Concept] OR "lumateperone" [Supplementary Concept] OR abilify[tiab] OR aripiprazole [tiab] OR aristada[tiab] OR brexpiprazole[tiab] OR brilaroxazine[tiab] OR caplyta[tiab] OR cariprazine[tiab] OR lumateperone[tiab] OR oxaripiprazole[tiab] OR reagila[tiab] OR rexulti[tiab] OR RP5063[tiab] OR vraylar[tiab] OR ((3rd generation[tiab] OR third generation[tiab]) AND antipsychotic*[tiab]) OR ( (D2[tiab] OR "D(2)"[tiab] OR D3[tiab] OR "D(3)"[tiab] OR dopamine receptor[tiab]) AND agonis*[tiab] AND partial[tiab]) | 6,906 |
| #1 | Search: "Suicide"[Mesh] OR "adverse effects" [Subheading:NoExp] OR "Safety"[Mesh] OR suicid*[tiab] OR adverse event*[tiab] OR adverse effect*[tiab] OR Safe*[tiab] | 3,268,822 |

**Cochrane Central Register of Controlled Trials (Wiley)**

| ID | Search | Hits |
|---|---|---|
| #1 | (suicid* OR (adverse NEXT event*) OR (adverse NEXT effect*) OR safe*):ti,ab,kw (Word variations have been searched) | 531323 |
| #2 | (abilify OR aripiprazole OR aristada OR brexpiprazole OR brilaroxazine OR caplyta OR cariprazine OR lumateperone OR oxaripiprazole OR reagila OR rexulti OR RP5063 OR vraylar OR (("3rd generation" OR "third generation") AND antipsychotic*) OR ( (D2 OR "D(2)" OR D3 OR "D(3)" OR "dopamine receptor") AND agonis* AND partial)):ti,ab,kw (Word variations have been searched) | 2301 |
| #3 | (schizo* OR psychoses OR psychosis OR psychotic OR "deficit syndrome"):ti,ab,kw (Word variations have been searched) | 27211 |
| #4 | #1 AND #2 AND #3 | 782 |
| #5 | #1 AND #2 AND #3 in Trials | 755 |

**PsycInfo (Ebsco)**

| # | Query | Results |
|---|---|---|
| S5 | S1 AND S2 AND S3 AND S4 | 442 |
| S4 | ( DE "Randomized Controlled Trials" OR DE "Treatment Effectiveness Evaluation" ) OR ( random* OR control* OR placebo ) | 1,031,916 |

| | | |
|---|---|---|
| S3 | ( DE "Acute Schizophrenia" OR DE "Affective Psychosis" OR DE "Brief Psychotic Disorder" OR DE "Capgras Syndrome" OR DE "Catatonic Schizophrenia" OR DE "Childhood Onset Psychosis" OR DE "Childhood Onset Schizophrenia" OR DE "Chronic Psychosis" OR DE "Delusional Disorder" OR DE "Paranoid Psychosis" OR DE "Paranoid Schizophrenia" OR DE "Postpartum Psychosis" OR DE "Process Schizophrenia" OR DE "Psychosis" OR DE "Reactive Psychosis" OR DE "Schizoid Personality Disorder" OR DE "Schizophrenia" OR DE "Schizophrenia (Disorganized Type)" OR DE "Schizophreniform Disorder" OR DE "Schizotypal Personality Disorder" OR DE "Undifferentiated Schizophrenia" ) OR ( schizo* OR psychoses OR psychosis OR psychotic OR "deficit syndrome" ) | 284,037 |
| S2 | DE "Aripiprazole" OR ( abilify OR aripiprazole OR aristada OR brexpiprazole OR brilaroxazine OR caplyta OR cariprazine OR lumateperone OR oxaripiprazole OR reagila OR rexulti OR RP5063 OR vraylar OR (("3rd generation" OR "third generation") AND antipsychotic*) OR ( (D2 OR "D(2)" OR D3 OR "D(3)" OR "dopamine receptor") AND agonis* AND partial) ) | 3,979 |
| S1 | ( DE "Attempted Suicide" OR DE "Military Suicide" OR DE "Suicidal Behavior" OR DE "Suicidal Ideation" OR DE "Suicidality" OR DE "Suicide" OR DE "Youth Suicide" OR DE "Patient Safety" OR DE "Safety" ) OR ( suicid* OR "adverse event*" OR "adverse effect*" OR Safe* ) | 273,127 |

### Scopus

| | Search Terms | Results |
|---|---|---|
| 5 | ( TITLE-ABS-KEY ( suicid* OR "adverse event*" OR "adverse effect*" OR safe* ) ) AND ( TITLE-ABS-KEY ( abilify OR aripiprazole OR aristada OR brexpiprazole OR brilaroxazine OR caplyta OR cariprazine OR lumateperone OR oxaripiprazole OR reagila OR rexulti OR rp5063 OR vraylar OR ( ( "3rd generation" OR "third generation" ) AND antipsychotic* ) OR ( ( d2 OR "D(2)" OR d3 OR "D(3)" OR "dopamine receptor" ) AND agonis* AND partial ) ) ) AND ( TITLE-ABS-KEY ( schizo* OR psychoses OR psychosis OR psychotic OR "deficit syndrome" ) ) AND ( ( INDEXTERMS ( "randomized controlled trial" OR "controlled clinical trial" OR "drug therapy" ) ) OR ( TITLE-ABS ( ( randomized OR randomised OR placebo OR randomly OR trial OR groups ) ) ) OR ( AUTHKEY ( random* OR rct OR trial ) ) ) | 1,780 document results |
| 4 | ( INDEXTERMS ( "randomized controlled trial" OR "controlled clinical trial" OR "drug therapy" ) ) OR ( TITLE-ABS ( ( randomized OR randomised OR placebo OR randomly OR trial OR groups ) ) ) OR ( AUTHKEY ( random* OR rct OR trial ) ) | 11,420,034 document results |
| 3 | TITLE-ABS-KEY ( schizo* OR psychoses OR psychosis OR psychotic OR "deficit syndrome" ) | 376,465 document results |

| | | |
|---|---|---|
| 2 | TITLE-ABS-KEY ( abilify OR aripiprazole OR aristada OR brexpiprazole OR brilaroxazine OR caplyta OR cariprazine OR lumateperone OR oxaripiprazole OR reagila OR rexulti OR rp5063 OR vraylar OR ( ( "3rd generation" OR "third generation" ) AND antipsychotic* ) OR ( ( d2 OR "D(2)" OR d3 OR "D(3)" OR "dopamine receptor" ) AND agonis* AND partial ) ) | 20,081 document results |
| 1 | TITLE-ABS-KEY ( suicid* OR "adverse event*" OR "adverse effect*" OR safe* ) | 3,593,236 document results |

**Europe PMC (preprints)**

(abilify OR aripiprazole OR aristada OR brexpiprazole OR brilaroxazine OR caplyta OR cariprazine OR lumateperone OR oxaripiprazole OR reagila OR rexulti OR rp5063 OR vraylar OR ( ( "3rd generation" OR "third generation" ) AND antipsychotic* ) OR ( ( d2 OR "D(2)" OR d3 OR "D(3)" OR "dopamine receptor" ) AND agonis* AND partial )) AND ( suicid* OR "adverse event*" OR "adverse effect*" OR safe* ) AND ( schizo* OR psychoses OR psychosis OR psychotic OR "deficit syndrome" ) AND (SRC:PPR)

39 results
